# The use of antibiotics commonly associated with antimicrobial resistance: a UK network cohort study using primary and hospital care data

**DOI:** 10.1101/2025.11.26.25340975

**Authors:** Elin Rowlands, Cecilia Campanile, Hiba Junaid, Jennifer C. E. Lane, Usama Rahman, Ben Eaton, Xavier L Griffin, Andy South, Ana Cavalcante, Duncan Cartner, Steve Harris, Ofran Almossawi, Lydia Briggs, Daniel Key, Prasiddha Khadka, Timothy Howcroft, Vishnu Vardhan Chandrabalan, Peter S Hall, Mahéva Vallet, Marta Pasikowska, Nicola Symmers, Colin McLean, Spyro Nita, Daniel Dedman, Zara Cuccu, Stelios Theophanous, Geoff Hall, Martí Català, Daniel Prieto Alhambra, Danielle Newby, Edward Burn

## Abstract

**Objectives:** To describe and compare patterns of use and potential indications of antibiotics in the WHO AWaRe classification Watch list in UK NHS community care and hospital care settings.

**Design:** Retrospective observational cohort study using routinely collected electronic health record data.

**Setting:** Community and hospital care in England and Scotland.

**Participants:** We included 12,139,211 records covering 33 different Watch antibiotics used in UK hospitals and community care settings.

**Main outcome measures:** Outcomes were incident prescription / dispensation records of each WHO AWaRe classification Watch list antibiotic between 2022 to last date of data capture for hospital care and from 2012 for community care. Conditions of interest captured 14 days before and after Watch list antibiotic use. Previous use of Access and other Watch list antibiotics were captured 14 days up to 1 day before each Watch list antibiotic.

**Results:** Our study found marked differences between community and hospital care in the use of Watch antibiotics. In community care, incidence trends for commonly used Watch antibiotics typically declined or remained stable between 2012 and 2024 (e.g., fusidate: from 2,471 to 438 per 100,000 person-years in CPRD Aurum; from 3,594 to 1,561 in DataLoch), with clear seasonality in their use and higher incidence between October and March. Watch antibiotic use in community care was typically linked to mild, common infections such as skin or respiratory tract infections. In hospital care, Watch antibiotic use was more often associated with clinically severe infections such as sepsis (e.g., vancomycin: from 12.2% of users in Leeds to 28.0% in Lancashire) and exacerbations of COPD (e.g., azithromycin: from 1.4% in GOSH to 65.7% in Lancashire). Prior use of Access and other Watch antibiotics was recorded in both community and hospital care, though this was generally observed in fewer than half of patients.

**Conclusions:** Overall, our findings suggest that Watch antibiotic use reflects the contrasting clinical contexts of community and hospital care, with milder infections predominating in the community and more severe infections in hospitals. However, regional and demographic variation indicates areas where prescribing practices could be refined. Ongoing surveillance and targeted stewardship interventions are essential to promote appropriate use and prevent resistance.

**Summary boxes:** *Section 1: What is already known on this topic:*

- Antibiotic prescribing / dispensing patterns can vary across care settings with most studies focusing on either community or hospital care in isolation.
- The WHO AWaRe classification highlights the need to monitor the use of Watch due to their higher potential to drive antimicrobial resistance.
- There is limited evidence across UK healthcare datasets examining real-world use of Watch list antibiotics and associated clinical indications.

*Section 2: What this study adds:*

- Variations in Watch list antibiotic prescribing / dispensing occur across community and hospital care settings.
- Conditions of interest of Watch list antibiotics patients were relatively consistent across general hospitals apart from paediatric hospital care and community care.

## Introduction

Bacterial infections are a major cause of morbidity and mortality worldwide (1). Antibiotics have been hugely successful in improving health outcomes and have significantly contributed to the global reduction in mortality in the under 5’s (2). While essential for treating bacterial infections, antibiotics pose a major public health challenge when misused, as this can accelerate the development of antimicrobial resistance (AMR) (3). AMR makes infections more difficult to treat and increases risks in medical procedures such as surgery and chemotherapy (4). In 2021, bacterial AMR was linked to 4.71 million deaths globally, including 1.14 million deaths directly attributed to resistant infections (5).

To help prevent AMR, the World Health Organization (WHO) introduced the AWaRe classification of antibiotics in 2017 as part of its global stewardship strategy. This classification categorised antibiotics into three groups: Access (low resistance potential), Watch (high resistance potential), and Reserve (last resort options) (6). The National Institute for Health and Care Excellence (NICE) guidelines generally recommend Access antibiotics as the first-line treatment for certain conditions, however Watch antibiotics are sometimes recommended as alternative first line therapy or second line therapy for common conditions such as urinary tract infections and respiratory tract infections (7, 8). Additionally, some infections require treatment with Watch antibiotics, for example Clostridioides difficile (9).

The use of Watch antibiotics should be carefully managed, as inappropriate use such as administering antibiotics without a clear clinical indication or without prior use of recommended first-line Access or Watch group antibiotics (if applicable) can contribute to the development and spread of AMR (10). However, little is known about the utilisation of Watch antibiotics in the United Kingdom (UK), particularly concerning patient characteristics and prescribing / dispensing practices. Careful management of Watch antibiotic use is needed to ensure they remain available when clinically necessary, while minimising exposure to prevent further resistance.

In this study, we aimed to better understand the trends, key characteristics and conditions of interest associated with Watch antibiotic use across community and hospital care in the UK. By providing insight into trends of Watch antibiotics use, our findings can support targeted stewardship strategies and inform policies to help mitigate the development and spread of AMR.

## Methods

### Study design

We conducted a network cohort study across the UK using routinely collected healthcare data mapped to the Observational Medical Outcomes Partnership Common Data Model (OMOP CDM) (11). The OMOP CDM allowed the study to be run by each site with common analytical code. Results were aggregated without sharing individual patient level data.

### Data sources

The study was performed across the recently established Health Data Research OMOP Network (HERON), which to date includes seven data partners covering different healthcare settings and geographies all mapped to the OMOP CDM (12). Five hospital databases were included: Barts Health NHS Trust (Barts), Great Ormond Street Hospital NHS Foundation Trust (GOSH), Lancashire Teaching Hospitals, Leeds Teaching Hospitals NHS Trust, and University College London Hospitals NHS Foundation Trust (UCLH), as well as two databases with community care data: the UK community care Clinical Practice Research Datalink (CPRD) Aurum and NHS Lothian (DataLoch) (**Table 1**).

**Table 1:** Database descriptions.

| Database | Person count | Release date | Description |
| --- | --- | --- | --- |
| Barts | 3,234,945 | 30/08/2024 | Captures data on patients from hospitals within the trust, which includes The Royal London Hospital, Newham General Hospital, Whipps Cross Hospital, Mile End Hospital, and St Bartholomew's Hospital. |
| CPRD Aurum | 50,370,041 | 20/09/2024 | The CPRD Aurum database contains longitudinal routinely-collected electronic health records (EHR data) from UK primary care practices using EMIS Web® general practice patient management software. |
| DataLoch | 1,753,685 | 05/08/2025 | The general population CDM contains person records drawn from the Lothian subsets of 4 national datasets: PHS Scottish Morbidity Records SMR00 (outpatient), SMR01 (inpatient), and SMR06 (cancer registry), and the PIS dataset for community prescribing information. These patients have been linked to historical coded data for hospitalisations, drug prescriptions, laboratory results, SIMD quintiles, GP data for BMI, alcohol consumption and smoking status, and deaths data from the National Records of Scotland. The dataset covers a geographical area in South East Scotland with a population of approximately 900k individuals. |
| GOSH | 203,386 | 05/11/2024 | GOSH is a specialist tertiary care paediatrics hospital in the UK receiving children with complex and/or rare conditions. A large proportion of the children that are seen at GOSH typically require long-term care and follow up at GOSH. This results in data collected during routine healthcare that are very rich in number of variables and detail. |
| IDRIL (Lancashire) | 1,533,222 | 02/01/2025 | Captures data from Chorley and South Ribble Hospital, Royal Preston Hospital, and The Specialist Mobility and Rehabilitation Centre |
| LTHT (Leeds) | 1,486,578 | 08/05/2025 | Captures hospital data from patients who received care within the trust. |
| UCLH | 1,389,095 | 08/05/2025 | Captures hospital data from patients who received care within the trust, which includes University College Hospital, Royal National Throat, Nose and Ear Hospital, National Hospital for Neurology and Neurosurgery, and the UCH Macmillan Cancer Centre. |

### Study Participants

The study period was from start of usable data to end of database capture. For CPRD Aurum and DataLoch, usable data was available from 2012, while for the remaining data partners this was from 2022. In the community care databases (CPRD Aurum and DataLoch), individuals were considered under observation from their first record (equivalent to GP registration in CPRD) to deregistration, death or end of the database. In hospital databases, individuals were under observation from the first recorded visit, condition, or drug exposure. Observation continued for up to 18 months following the most recent record. Overlapping observations were merged in single observation episodes (e.g. two records separated by less than 18 months lead to a single observation period from the first record to 18 months after the second record). Observation stopped at death or end of database. In GOSH, which is a paediatric database, individuals were no longer considered under observation once they reached 18 years of age. For hospital databases, only Watch antibiotics used during an inpatient or Accident & Emergency (A&E) visit were included to capture Watch antibiotics use during acute hospital care.

### Primary outcomes

The main outcome of interest was prescription / dispensation of each antibiotic from the Watch category of the WHO AWaRe classification (2023). All drug concepts that included a Watch antibiotic, whether alone or in combination with other active ingredients, were included. Only antibiotics with at least 100 patient records during the study period were included. In total, we included 134 Watch antibiotics (**Table S1**).

Individuals could contribute multiple records of Watch antibiotic use, provided there was a wash-out period of at least 30 days between exposures. Records within seven days of each other were merged.

### Statistical methods

Descriptive statistics were used to summarise characteristics for patient records for each Watch antibiotic for each database. Use of Watch and Access antibiotics prior to initiation of each Watch antibiotic and defined conditions of interest were also summarised. Median (IQR) and Mean (SD) were used for continuous variables and counts and percentages used for categorical variables. Characterisation was performed using the CohortCharacteristics R package (13).

Overall and quarterly crude incidence rates (IR) for each Watch antibiotic were calculated from 2022 to the end of available data for each database. For incidence, the number of events, the observed time at risk, and the incidence rate per 100,000 person years were summarised along with 95% confidence intervals (95% CI). We calculated 95% confidence intervals of the estimated rates using the Poisson distribution. We allowed for repetitive events. Incidence rates were calculated overall and stratified by age group and sex. Incidence was estimated using the IncidencePrevalence R package (14).

Counts smaller than five were suppressed across all databases to prevent re-identification. For Leeds database counts smaller than six were suppressed, and for DataLoch counts smaller than ten were suppressed. All our analytical code is available in a public GitHub repository: https://github.com/heron-uk/HDRUK-01-001-Antibiotics.

### Patient and Public Involvement

This research question was chosen in collaboration with key national stakeholders, including HDRUK, HDR Alliance, the NHS, NICE, MHRA, and UKHSA including patients and public.

## Results

### Overview

A total of 433,368 and 13,423,216 records of Watch antibiotic use were found in hospital and community care settings, respectively. Across all databases, we had results for 33 different Watch antibiotics. All study results can be found in the accompanying Shiny app (https://dpa-pde-oxford.shinyapps.io/heronAntibioticsStudy/).

In CPRD Aurum, the most used Watch antibiotic between 2012 and 2025 was neomycin with 4,477,598 records (**Table 2**). The most used Watch antibiotic in DataLoch between 2012 and 2025 was clarithromycin, with 394,394 records. The combination of piperacillin and tazobactam was the most frequently used Watch antibiotic in Barts (n = 19,976), Leeds (n = 26,365), and GOSH (n = 4,227) between 2022 and 2025, while cefuroxime was most frequent in Lancashire (n = 10,801) and UCLH (n = 44,228) (Table 2). As the results for piperacillin and tazobactam were nearly identical, only the results for piperacillin are presented, but it is labeled as piperacillin/tazobactam. Patient demographics for the top five Watch antibiotic for each database, including age and sex, can be found in **Tables S4 – S10**.

**Table 2:** Five most commonly used Watch antibiotics, ranked by number of records (n), in each database. Antibiotics in top five among at least five databases are shown in bold.

| Rank | CPRD Aurum | Barts | DataLoch | GOSH | Lancashire | Leeds | UCLH |
| --- | --- | --- | --- | --- | --- | --- | --- |
| 1 | Neomycin (4,477,598) | **Piperacillin / Tazobactam (19,976)** | **Clarithromycin (394,394)** | **Piperacillin / Tazobactam (4,227)** | Cefuroxime (10,801) | **Piperacillin / Tazobactam (26,365)** | Cefuroxime (44,228) |
| 2 | Fusidate (2,029,304) | **Clarithromycin (11,794)** | Fusidate (390,456) | Teicoplanin (2,025) | **Clarithromycin (9,870)** | **Clarithromycin (11,567)** | **Ciprofloxacin (19,251)** |
| 3 | **Ciprofloxacin (1,561,067)** | **Ciprofloxacin (11,359)** | Neomycin (353,001) | Vancomycin (1,757) | **Piperacillin / Tazobactam (6,642)** | Cefuroxime (11,270) | **Piperacillin / Tazobactam (10,450)** |
| 4 | Erythromycin (1,295,225) | Ceftriaxone (10,841) | Oxytetracycline (178,584) | **Ciprofloxacin (1,571)** | **Ciprofloxacin (2,400)** | **Ciprofloxacin (10,043)** | **Clarithromycin (9,842)** |
| 5 | Clarithromycin (769,070) | Meropenem (7,417) | Erythromycin (136,037) | Meropenem (1,527) | Levofloxacin (2,314) | Teicoplanin (9,608) | Teicoplanin (5,547) |

### Incidence rates

In hospital care, quarterly crude IR trends of the most commonly used Watch antibiotics varied across sites, but stable short-term trends were also observed (**Figure 1**). For example, piperacillin/tazobactam use remained relatively stable across all hospitals ranging from 157.23 to 4,393 per 100,000 person years in Lancashire and Leeds respectively with GOSH showing a decline in IRs from 1,786 [95% CI 1,601 - 1,987] to 1,349 [1,198 - 1,514] per 100,000 person years between 2022 and Q3 2023 before increasing. Excluding GOSH, clarithromycin IRs showed large variability over time across all hospitals apart from UCLH where trends remained stable with IR from 452.9 [415.6 - 492.5] in Q1 2022 to 498.7 [462.6 - 536.9] in Q3 2024. Other notable trends include for Barts, ceftriaxone showing stable IRs between Q1 2022 and Q2 2023 before a sharp increase with stability for the rest of the study period. Additionally, cefuroxime trends showed stability with gradual increases in Leeds and UCLH whereas for Lancashire stable IRs were seen either side of a large drop in IRs between 2023 to 2024.

**Figure 1:**
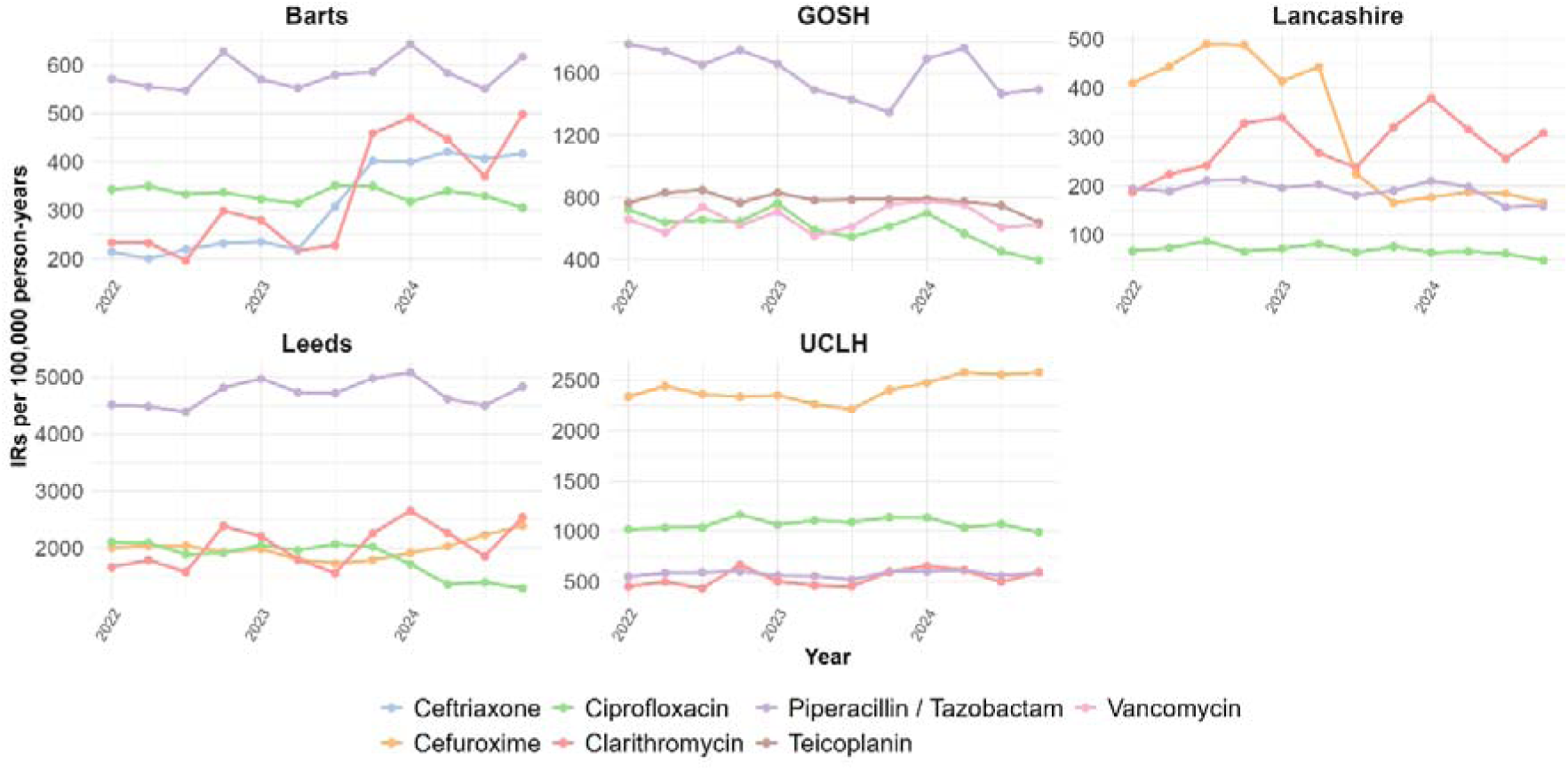
Quarterly crude incidence rates of the five most used Watch antibiotics across hospital databases.

In community care, neomycin had the highest incidence across the study period for CPRD Aurum (**Figure 2**) showing stable IRs from 2,183 [2,168 - 2,199] per 100,000 person years in Q1 2012 to 2,090 [2,074 - 2,106] in Q4 2024. Incidence of neomycin in DataLoch showed a decline over the study period (2,464 in Q1 2012 to 1,744 in Q3 2024). Clarithromycin had the highest incidence across the study period for DataLoch, however the IRs declined significantly from 4,883 [4,799 – 4,968] in Q1 2012 to 1,510 [1,471 – 1,551] in Q4 2024. In contrast, IRs of clarithromycin in CPRD Aurum remained relatively stable (438 in 2012 to 395 in 2024).

**Figure 2:**
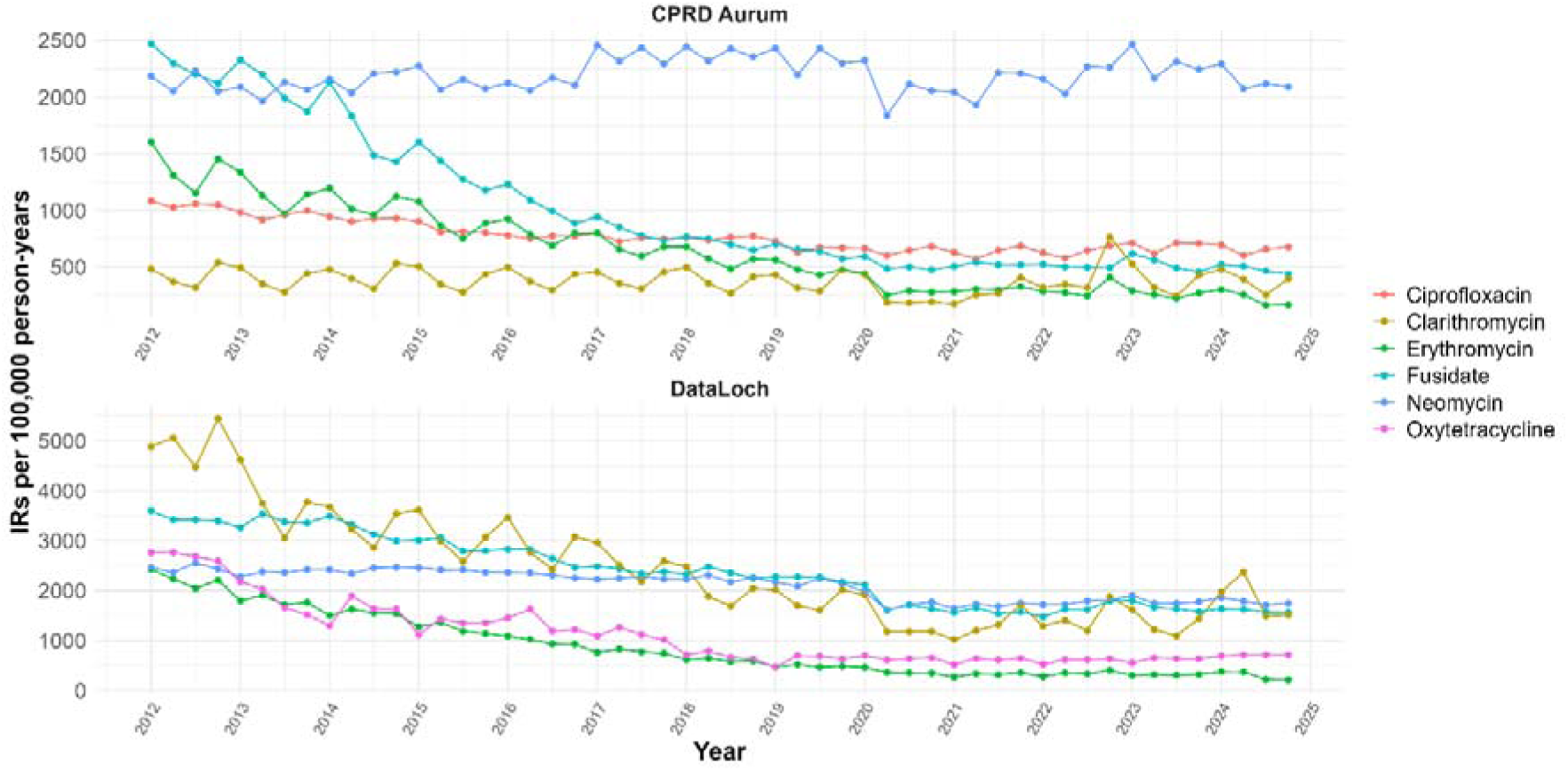
Quarterly crude incidence rates of the five most used Watch antibiotics across community care databases.

Fusidate and erythromycin showed decreasing IRs between Q1 2012 and Q1 2020 in both CPRD Aurum (from 2,471 [2,454–2,487] to 593 [585–600], and from 1,604 [1,591–1,617] to 440 [434–447] per 100,000 person-years, respectively) and DataLoch (from 3,594 [3,522–3,668] to 2,112 [2,063–2,161], and from 2,428 [2,369–2,488] to 463 [441–487] per 100,000 person-years, respectively). From 2020 onwards, IRs remained stable until the end of the study period. Across all antibiotics, IRs displayed seasonality, typically peaking in the third and fourth quarters of each year.

There were some differences in IRs between males and females for several of the top Watch antibiotics in both hospital and community care databases (**Figure S1-S7**). While both sexes followed similar trends, incidence was higher in females for neomycin, erythromycin, and fusidate in community care. There were no sex differences for clarithromycin in CPRD Aurum, however incidence was higher in females for clarithromycin in DataLoch. Additionally, there were no sex differences for ciprofloxacin in CPRD Aurum or for oxytetracycline in DataLoch. In hospital care, there were either no sex differences, or males had higher IRs, with some exceptions, for example cefuroxime showed higher IRs in females at Leeds and UCLH.

There were also some differences in IRs between different age groups in community care (**Figures S8-S14**). For both CPRD Aurum and DataLoch, incidence of neomycin was highest in patients ≥65, and incidence of erythromycin was highest in children (patients under 18). However, for CPRD Aurum incidence of clarithromycin use was highest in children, while in DataLoch it was highest in patients over 65. However, all age groups typically followed the same trends over the study period.

In all hospitals except GOSH, incidence of the top five antibiotics is typically highest in patients ≥65, with some exceptions. In Leeds, incidence of cefuroxime was highest in patients 18-64 (overall incidence: 2,819 (2,759 – 2,880), followed by patients under 18 (1,464 (1,372 – 1,560)). However in Lancashire and UCLH overall incidence was highest in patients ≥65 (584 (570 - 599) and 3,051 (2,999 – 3,104) respectively). Additionally, no significant difference was found in overall incidence of teicoplanin use in Leeds between the three age groups (0-17: 1,691 (1,592 – 1,794), 18-64: 1,414 (1,372 – 1,457), ≥65: 1,908 (1,851 – 1,965)), however in UCLH overall incidence was highest in the ≥65 group (463 (443-484)), and similar in the 0-17 and 18-64 groups (212 (192 – 232) and 203 (196 – 210) respectively).

### Prior use of Access and Watch antibiotics

Prior use of Access antibiotics within 14 days before Watch antibiotics showed some variation across databases although there were some similarities (**Figure 3**). In the hospital care databases, amoxicillin was one of the most frequently prescribed Access antibiotics prior to the Watch antibiotic piperacillin/tazobactam ranging from 15.3% in Leeds to 51.2% in Barts. In community care, amoxicillin was often the only Access antibiotic prescribed in more than 5% of cases before Watch antibiotic use. Distinct patterns were seen for GOSH, where amikacin was among the most prescribed prior Access antibiotics prior to Watch antibiotics ranging from 11.9% for piperacillin/tazobactam and 45.6% for vancomycin. At UCLH, trimethoprim and sulfamethoxazole were prescribed prior to Watch antibiotic, a pattern not seen in other hospital care databases, whereas gentamicin, was prescribed prior to the top Watch antibiotics for Lancashire. For DataLoch, prior use of Access antibiotics was identified in fewer than 5% of users of the five most common Watch antibiotics, and therefore wasn’t presented in Figure 3.

**Figure 3:**
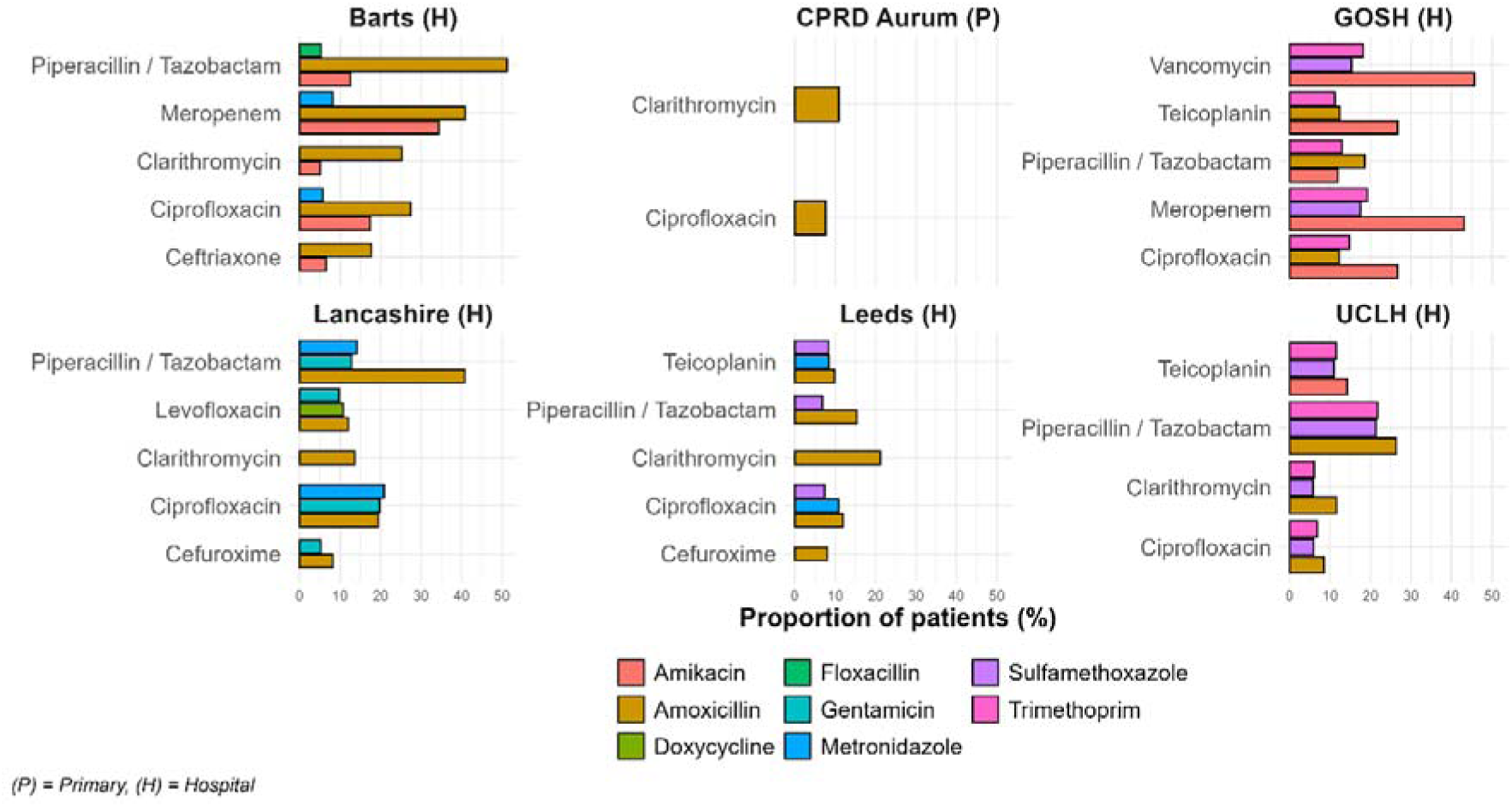
Use of Access antibiotics 14 days prior to use of a top five Watch antibiotic, stratified by database. Only includes Access antibiotics recorded in at least 5% of users.

Prior use of other Watch antibiotics within 14 days before Watch antibiotics also showed variation across databases (**Figure 4**). Piperacillin/tazobactam was commonly given prior to other Watch antibiotics in hospital care. For example, in Barts and GOSH approximately a third of patients that received meropenem had a record of piperacillin/tazobactam use in the 14 days prior (33.0% and 33.5% respectively). Cefuroxime was also used before certain Watch antibiotics for Lancashire, Leeds and UCLH, including ciprofloxacin (18.5%, 10.1%, and 7.8%, respectively). For both CPRD Aurum and DataLoch, prior use of other Watch antibiotics was identified in fewer than 5% of users of the five most common Watch antibiotics, therefore were not presented in Figure 4.

**Figure 4:**
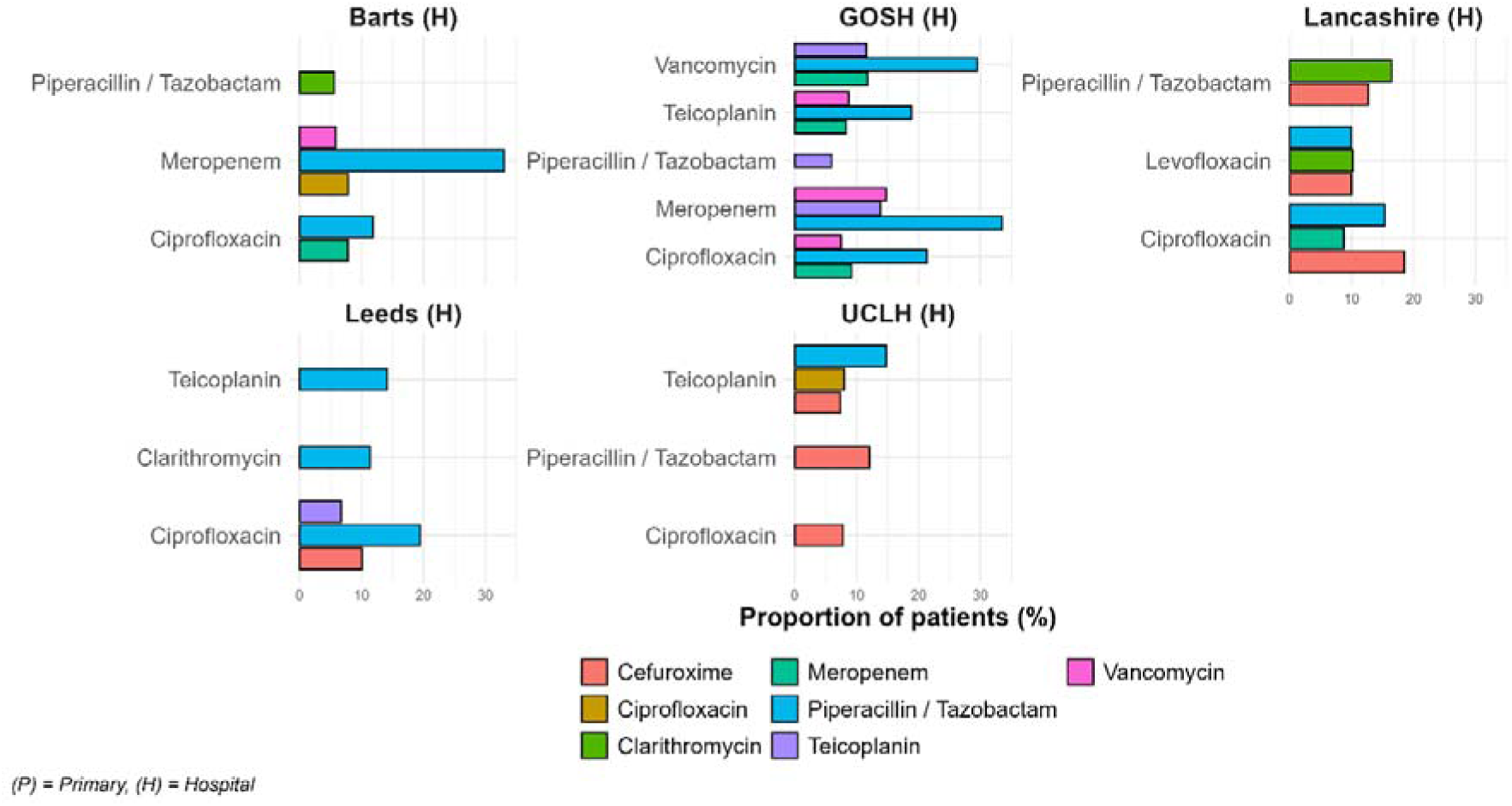
Use of other Watch antibiotics 14 days prior to use of a top five Watch antibiotic, stratified by database. Only includes Watch antibiotics recorded in at least 5% of users.

### Conditions of interest related to antibiotic treatment indications

There were a wide range of conditions of interest recorded alongside Watch antibiotic use, with differences between community and hospital care, as well as some differences between hospitals (**Figure 5**).

**Figure 5:**
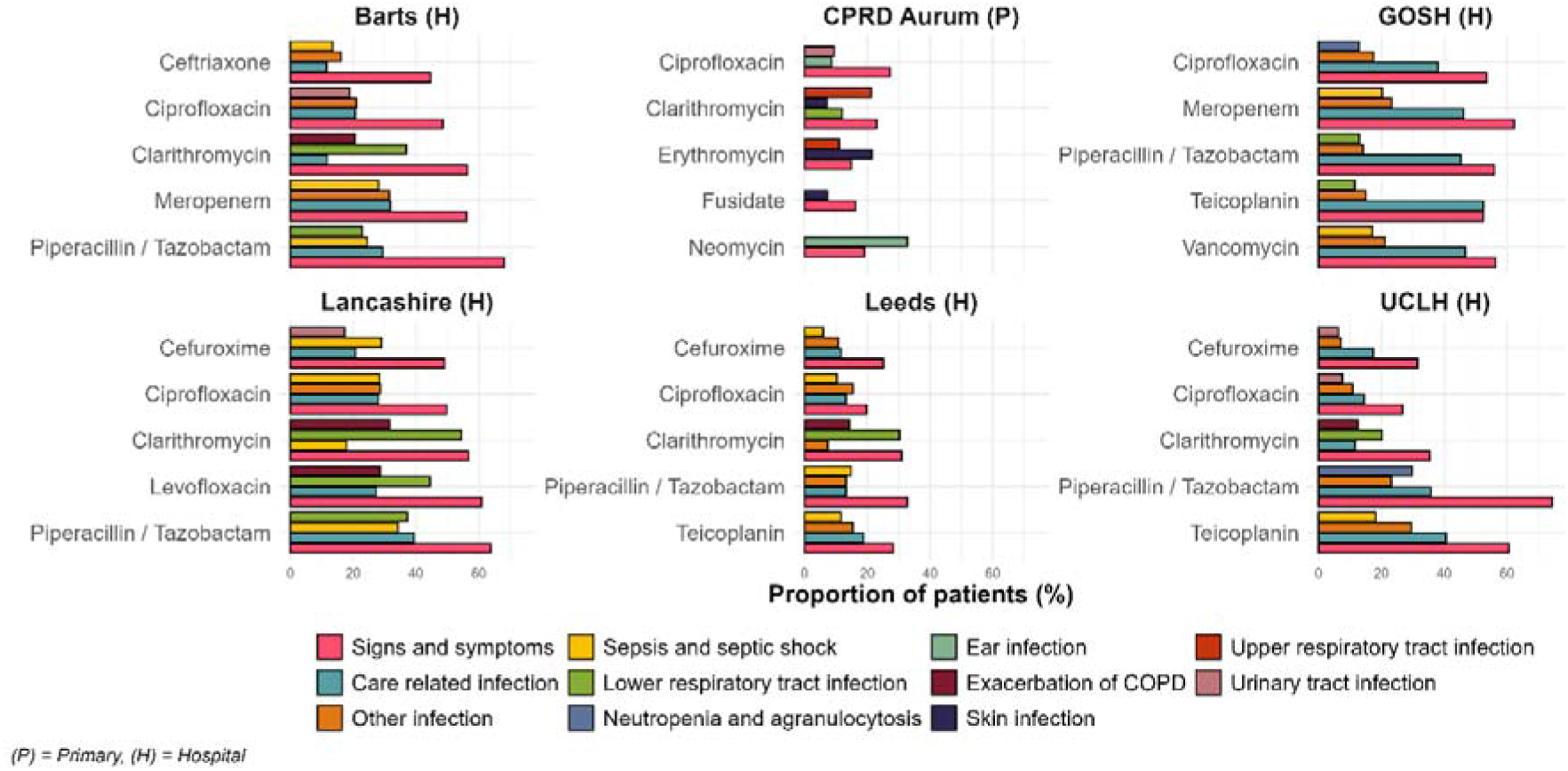
The four most frequent conditions of interest recorded in patients in the 14 days prior and 14 days after top five Watch antibiotics across databases. Only includes conditions recorded in at least 5% of users.

In all hospital care databases except GOSH, lower respiratory tract infections were commonly associated with clarithromycin use, ranging from 11.7% in UCLH to 54.4% in Lancashire with exacerbations of COPD also recorded around the time of use (12.6% - 31.7%). For meropenem, Barts and GOSH showed similar patterns, with sepsis and septic shock (28.1% and 20.3%), care-related infections (31.8% and 46.2%), and other infections (31.5% and 23.3%) recorded. For ciprofloxacin, sepsis and septic shock were recorded in Lancashire and Leeds (28.4% and 10.3%) whereas Barts and UCLH recorded urinary tract infections (18.9% and 7.6% respectively) on top of care related infections and other infections. For GOSH, care related infections were the most frequently recorded conditions, ranging from 38.0% for ciprofloxacin and 52.5% for teicoplanin. Care related infections were also commonly recorded among the top Watch antibiotics for UCLH, ranging from 11.6% for clarithromycin to 40.6% for teicoplanin. In community care, CPRD Aurum showed ear infections recorded alongside neomycin (32.7%) while fusidate, erythromycin and clarithromycin were predominately recorded alongside skin infections.

## Discussion

### Principal findings

We analysed over 13.8 million records of Watch antibiotic use from seven UK databases. There was strong consistency across hospitals, with piperacillin/tazobactam appearing in the top five antibiotics at every site, although the associated conditions of interest varied: care-related infections were most common at Barts, GOSH, Lancashire, and UCLH, while sepsis and septic shock were the most common at Leeds. Ciprofloxacin also featured in the top five across all hospitals, with variation in associated conditions: it was linked to ‘other’ infections (site not specified) at Barts, Lancashire, and Leeds, and to care-related infections at GOSH and UCLH. A high proportion of ‘signs and symptoms’ were observed across all top five Watch antibiotics in hospital care.

Quarterly incidence rates of Watch antibiotic use and patient characteristics varied by hospital, however higher incidence of Watch antibiotic use in male patients was consistently observed across all hospitals, in contrast to community care, where use was generally higher among female patients. In both community and hospital care, many patients using Watch antibiotics had prior use of Access and/or other Watch antibiotics, although this was typically observed in fewer than half of patients.

Although there was minimal overlap in the top five Watch antibiotics between community and hospital care, four of the top five antibiotics were the same in CPRD Aurum and DataLoch. Both databases also showed similar trends, with most Watch antibiotics declining over the study period, including erythromycin and fusidate. Patient characteristics were also broadly consistent.

### Comparison with other studies

Our findings align with previous UK and international research, demonstrating variability in antibiotic use across care settings and regions (15–19). Variation in hospital care use is likely driven by stewardship protocols, local governance, and hospital-specific policies with some centres managing more resistant or complex infections (20). Regional microbiological variation, including differences in bacterial strains and resistance may necessitate different prescribing / dispensing practices (21, 22). Socio-demographics, health-seeking behaviour, comorbidity profiles, and differences between adult and paediatric populations further impacting use patterns (23, 24).

Differences between community and hospital care settings reflect the role of care setting in shaping prescribing / dispensing and stewardship priorities in the NHS. In community care, most uses relate to common and milder infections such as upper respiratory tract infections, often treated with Access list antibiotics (25). In contrast, Watch antibiotics were more frequently used in hospital care to treat severe infections, surgical prophylaxis, or for patients requiring broader-spectrum coverage (10, 26).

The use of Watch list antibiotics should ideally follow appropriate first-line treatment, which are typically Access group antibiotics or in some cases other Watch antibiotics. We found that many patients had used Access antibiotics prior to initiation of a Watch antibiotic, suggesting escalation in line with clinical guidelines (10). However, prior use of Access antibiotics was typically recorded in less than 50% of patients, suggesting Watch antibiotics were also commonly used as first line treatment, possibly due to clinically assessed severity and/or bacterial resistance (27).

We observed higher Watch antibiotic use in female patients in community care, aligning with previous studies (28, 29). This may reflect healthcare-seeking behaviours, with females being more likely to seek community care treatment (30). We also observed higher Watch antibiotic use in male patients in hospital care, which further supports this interpretation, as not seeking healthcare promptly could lead to more severe infections that require hospital-based treatment.

### Strengths and weaknesses of the study

A major strength of this study is the large-scale encompassing over 13.8 million patient records across multiple databases and care settings. This comprehensive approach enhances generalisability and offers a more complete picture of antibiotic use in the UK. Additionally, this study provides unique insights that cannot be obtained using national data, as hospital-level information is not routinely collected. By using OMOP, it was possible to directly access data from care providers and enable distributed analytics without transferring patient-level data.

This study has several limitations. First, the conditions used to define conditions of interest were based on data-driven characterisation, which may have missed rare conditions or introduced misclassification. Second, the 14-day window may have missed some conditions, for example if use was recorded on the date of discharge. Third, our analysis used the WHO 2023 AWaRe list, which may not fully reflect UK-specific adaptations (31). Fourth, because community and hospital care data were generally not linked, we could not access prior community care records for hospital patients, and we expect some overlap across databases. Finally, due to data limitations, we analysed antibiotics only at the ingredient level, without information on route or dose. This may affect interpretation, as different routes of administration for the same antibiotic can fall into different AWaRe categories.

### Implications for clinicians and policy makers

Understanding variations in antibiotic use, particularly those on the Watch list across care settings across the UK is critical for shaping targeted antimicrobial stewardship interventions, particularly with growing rates of AMR (32). Our findings suggest that although there is some consistency in trends of use, clinical conditions of interest and in the prior use of Access and/or Watch antibiotics in line with guidelines, there is variability across databases, particularly within hospital care. This highlights opportunities to improve standardisation of prescribing / dispensing practices and strengthen adherence to clinical guidelines. Unnecessary use due to lack of appropriate indication is associated with almost a third of global antibiotic consumption (33). Therefore, our results give crucial insight into current use of Watch antibiotics in the UK and underscores the need for clear guidelines to ensure appropriate use in both community and hospital care. For clinicians, this study highlights increased awareness of resistance trends and the importance of accurate recording of infections or conditions that require antibiotic treatment. For policymakers, the study highlights the need for integrated stewardship programmes that span both community and hospital care.

### Unanswered questions and future research

Further research is needed to explore the underlying causes of observed variation in Watch antibiotic use, including regional factors, clinician preferences, and specific patient characteristics and demographics as well as changes in global and national watchlists (34). In addition, further research is needed to understand regional differences in dose and route of administration, and the impact this may have on AMR. Understanding how local governance structures, recommendations and costings influence antibiotic selection across the AWaRe classification in different regions could also help inform more effective stewardship strategies. Furthermore, qualitative research may also help elucidate decision-making processes that are not captured in structured data. Future studies should also incorporate microbiological data to assess appropriateness of antibiotic use and resistance patterns (35).

## Ethics approval

This work uses data provided by patients and collected by the NHS as part of their care and support. Informed consent of individual patients was not required as anonymised information was obtained from medical records. The ethical approval for each database is as follows:

### Clinical Practice Research Datalink (CPRD)

Protocol for this research was approved by the independent scientific advisory committee for Medicine and Healthcare Products Regulatory Agency database research (protocol No 24_004673)

### Barts Health

The Project was registered with the Clinical Effectiveness Unit at Barts Health NHS Trust, and the data access request was approved by the Barts Health Data Access Committee (DAC035).

### Leeds Teaching Hospitals

The protocol for this study was approved by the Trust’s Data Access Committee (approval ref: LTH25002 DR1)

**DataLoch:** approval was granted by the NHS Lothian DataLoch governance process that operates within NHS Lothian with authority delegated from the NHS Lothian Caldicott Guardian, NHS Research Ethics Service and the NHS Lothian Research and Development department (ACCORD).; DataLoch service for research / REC 22/NS/0093 / IRAS 317626.

### University College London Hospital (UCLH)

The study was deemed exempt from NHS Research Ethics Committee review as there is no change to treatment or services or any study randomisation of patients into different treatment groups. It was considered a Service Evaluation according to the NHS Health Research Authority decision tool (http://www.hra-decisiontools.org.uk/research/).

### Great Ormond Street Hospital (GOSH)

This research was carried out under our general approvals for the use of routinely collected data in research (21SH28, IRAS 301663) and participation in HERON (24SH30). The specific data release was approved by the Data Partnerships Committee as per the conditions of the latter.

### Lancashire Teaching Hospitals NHS Foundation Trust

The OMOP Database at LTH has been approved by the NHS Research Ethics Committee for real world evidence studies (REC Reference 24/NI/0001 IRAS Reference 315234)

## Data availability statement

Patient level data cannot be shared without approval from data custodians owing to local information governance and data protection regulations. Analytical code for the study is freely available in a GitHub repository (https://github.com/heron-uk/HDRUK-01-001-Antibiotics). All study results can be found in the accompanying Shiny app (https://dpa-pde-oxford.shinyapps.io/heronAntibioticsStudy/).

## Acknowledgments

We thank members of the review panel, Alison Cave (MHRA), Chris Russell, Andy Payne and Phil Waywell (NHS England), Pall Jonsson and Shaun Rowark (NICE), Yvonne Adebola (useMYdata), and Joshua Symons and Gavin Dabrera (UKHSA). Additionally, we thank Alex Knight, Paola Quattroni, David Seymour and Uwaye Ideh (HDRUK) for their collaboration in establishing the HERON-UK network.

We also thank members of key national stakeholders, including HDR UK, the HDR Alliance, the NHS, NICE, MHRA, UKHSA, and patient representatives, for their valuable input on the initial study concepts.

## Transparency statement

The corresponding author (the manuscript’s guarantor) affirms that the manuscript is an honest, accurate, and transparent account of the study being reported; that no important aspects of the study have been omitted; and that any discrepancies from the study as planned (and, if relevant, registered) have been explained.

## Contributor and guarantor information

EB and D P-A conceived the study and contributed to the study design.

ER, DN and EB wrote and reviewed the analytical code with ST, LB, TH, DD, HJ, AS and NS conducting statistical analyses at each data partner site.

ER, DN, DP-A and EB interpreted the results and wrote the first draft of the manuscript. All authors reviewed the manuscript, approved the final version, and had final responsibility for the decision to submit for publication. EB and DN are guarantors. The corresponding author attests that all listed authors meet authorship criteria and that no others meeting the criteria have been omitted.

## Copyright/license for publication

The Corresponding Author grants on behalf of all authors, a worldwide licence to the Publishers and its licensees in perpetuity, in all forms, formats and media (whether known now or created in the future), to i) publish, reproduce, distribute, display and store the Contribution, ii) translate the Contribution into other languages, create adaptations, reprints, include within collections and create summaries, extracts and/or, abstracts of the Contribution, iii) create any other derivative work(s) based on the Contribution, iv) to exploit all subsidiary rights in the Contribution, v) the inclusion of electronic links from the Contribution to third party material where-ever it may be located; and, vi) licence any third party to do any or all of the above.

## Dissemination to participants and related patient and public communities

We will disseminate a lay summary of our findings through our Twitter and other social media accounts.

