## Supplementary Information for "The use of antibiotics commonly associated with antimicrobial resistance: a UK network cohort study using primary and hospital care data"

### Table S1: Antibiotic from the Watch category of the WHO AWaRe classification (2023)

| **Concept ID** | **Ingredient Name** | **Vocabulary** |
| --- | --- | --- |
| 19101402 | arbekacin | RxNorm |
| 35198192 | aspoxicillin hydrate | RxNorm Extension |
| 1734104 | azithromycin | RxNorm |
| 19015123 | azlocillin | RxNorm |
| 36849346 | BEKANAMYCIN | RxNorm Extension |
| 35198093 | biapenem | RxNorm Extension |
| 1740546 | carbenicillin | RxNorm |
| 1768849 | cefaclor | RxNorm |
| 19070174 | cefamandole | RxNorm |
| 43009082 | cefbuperazone sodium | RxNorm Extension |
| 43009044 | cefcapene pivoxil hydrochloride hydrate | RxNorm Extension |
| 1796458 | cefdinir | RxNorm |
| 1747005 | cefditoren | RxNorm |
| 1748975 | cefepime | RxNorm |
| 19028241 | cefetamet | RxNorm |
| 1796435 | cefixime | RxNorm |
| 40798704 | Cefmenoxime | RxNorm Extension |
| 19072255 | cefmetazole | RxNorm |
| 43008993 | cefminox sodium | RxNorm Extension |
| 19028286 | cefodizime | RxNorm |
| 19072857 | cefonicid | RxNorm |
| 1773402 | cefoperazone | RxNorm |
| 19028288 | ceforanide | RxNorm |
| 36848891 | CEFOSELIS | RxNorm Extension |
| 1774470 | cefotaxime | RxNorm |
| 1774932 | cefotetan | RxNorm |
| 19051271 | cefotiam | RxNorm |
| 1775741 | cefoxitin | RxNorm |
| 35197975 | cefozopran hydrochloride | RxNorm Extension |
| 43009045 | cefpiramide sodium | RxNorm Extension |
| 19001904 | cefpirome | RxNorm |
| 1749008 | cefpodoxime | RxNorm |
| 1738366 | cefprozil | RxNorm |
| 19051345 | cefsulodin | RxNorm |
| 1776684 | ceftazidime | RxNorm |
| 35198137 | cefteram pivoxil | RxNorm Extension |
| 1749083 | ceftibuten | RxNorm |
| 1777254 | ceftizoxime | RxNorm |
| 1777806 | ceftriaxone | RxNorm |
| 1778162 | cefuroxime | RxNorm |
| 19095043 | chlortetracycline | RxNorm |
| 1797258 | cilastatin | RxNorm |
| 997496 | cinoxacin | RxNorm |
| 1797513 | ciprofloxacin | RxNorm |
| 1750500 | clarithromycin | RxNorm |
| 19047240 | clofoctol | RxNorm |
| 19047265 | clomocycline | RxNorm |
| 1592954 | delafloxacin | RxNorm |
| 1714527 | demeclocycline | RxNorm |
| 19023508 | dibekacin | RxNorm |
| 1790024 | dirithromycin | RxNorm |
| 1713905 | doripenem | RxNorm |
| 1743222 | enoxacin | RxNorm |
| 1717963 | ertapenem | RxNorm |
| 1746940 | erythromycin | RxNorm |
| 40239985 | fidaxomicin | RxNorm |
| 19050750 | fleroxacin | RxNorm |
| 43009087 | flomoxef sodium | RxNorm |
| 19064329 | flumequine | RxNorm |
| 36852196 | FLURITHROMYCIN | RxNorm Extension |
| 956653 | fosfomycin | RxNorm |
| 19010400 | fusidate | RxNorm |
| 35197938 | garenoxacin mesilate hydrate | RxNorm Extension |
| 1789276 | gatifloxacin | RxNorm |
| 1716721 | gemifloxacin | RxNorm |
| 1747032 | grepafloxacin | RxNorm |
| 1778262 | imipenem | RxNorm |
| 43009022 | isepamicin sulfate | RxNorm Extension |
| 19123240 | josamycin | RxNorm |
| 1784749 | kanamycin | RxNorm |
| 35834909 | lascufloxacin hydrochloride | RxNorm Extension |
| 1742253 | levofloxacin | RxNorm |
| 1790692 | lincomycin | RxNorm |
| 1707800 | lomefloxacin | RxNorm |
| 1708100 | loracarbef | RxNorm |
| 19092353 | lymecycline | RxNorm |
| 1709170 | meropenem | RxNorm |
| 19003644 | methacycline | RxNorm |
| 19007701 | mezlocillin | RxNorm |
| 19072089 | micronomicin | RxNorm |
| 19072122 | midecamycin | RxNorm |
| 1708880 | minocycline | RxNorm |
| 19009138 | miocamycin | RxNorm |
| 19126622 | moxalactam | RxNorm |
| 1716903 | moxifloxacin | RxNorm |
| 36878831 | nadifloxacin | RxNorm Extension |
| 36860386 | NEMONOXACIN | RxNorm Extension |
| 915981 | Neomycin | RxNorm |
| 19017585 | netilmicin | RxNorm |
| 1721543 | norfloxacin | RxNorm |
| 923081 | Ofloxacin | RxNorm |
| 19023254 | oleandomycin | RxNorm |
| 19129642 | oxolinic acid | RxNorm |
| 925952 | oxytetracycline | RxNorm |
| 35197853 | panipenem | RxNorm Extension |
| 35198003 | pazufloxacin mesilate | RxNorm Extension |
| 19027679 | pefloxacin | RxNorm |
| 19088795 | phenethicillin | RxNorm |
| 19010564 | pipemidate | RxNorm |
| 1746114 | piperacillin | RxNorm |
| 40799027 | Piromidic Acid | RxNorm Extension |
| 19125201 | pristinamycin | RxNorm |
| 35197897 | prulifloxacin | RxNorm Extension |
| 43009067 | ribostamycin sulfate | RxNorm Extension |
| 1777417 | rifabutin | RxNorm |
| 1763204 | rifampin | RxNorm |
| 19035924 | rifamycin SV | RxNorm |
| 1735947 | rifaximin | RxNorm |
| 35198144 | rokitamycin | RxNorm Extension |
| 19136024 | rolitetracycline | RxNorm |
| 19036545 | rosoxacin | RxNorm |
| 19063874 | roxithromycin | RxNorm |
| 35884386 | Rufloxacin | RxNorm Extension |
| 35200881 | sarecycline | RxNorm |
| 19136044 | sisomicin | RxNorm |
| 35198165 | sitafloxacin hydrate | RxNorm Extension |
| 36855357 | SOLITHROMYCIN | RxNorm Extension |
| 1733765 | sparfloxacin | RxNorm |
| 19070251 | spiramycin | RxNorm |
| 1836191 | streptomycin | RxNorm |
| 19136210 | streptozocin | RxNorm |
| 19000817 | sulbenicillin | RxNorm |
| 1741122 | tazobactam | RxNorm |
| 35198145 | tebipenem pivoxil | RxNorm Extension |
| 19078399 | teicoplanin | RxNorm |
| 1702911 | telithromycin | RxNorm |
| 19041153 | temafloxacin | RxNorm |
| 19100438 | temocillin | RxNorm |
| 1702364 | ticarcillin | RxNorm |
| 902722 | Tobramycin | RxNorm |
| 36857682 | TOSUFLOXACIN | RxNorm Extension |
| 19006043 | troleandomycin | RxNorm |
| 1712549 | trovafloxacin | RxNorm |
| 1707687 | vancomycin | RxNorm |

### Table S2: Antibiotic from the Access category of the WHO AWaRe classification (2023)

| **Concept ID** | **Ingredient Name** | **Vocabulary** |
| --- | --- | --- |
| 19123877 | amdinocillin | RxNorm |
| 19088223 | amdinocillin pivoxil | RxNorm |
| 1790868 | amikacin | RxNorm |
| 1713332 | amoxicillin | RxNorm |
| 1717327 | ampicillin | RxNorm |
| 19018516 | azidocillin | RxNorm |
| 1734205 | bacampicillin | RxNorm |
| 19018742 | brodimoprim | RxNorm |
| 40798709 | cefacetrile | RxNorm Extension |
| 1769535 | cefadroxil | RxNorm |
| 19070680 | cefatrizine | RxNorm |
| 40798700 | cefazedone | RxNorm Extension |
| 1771162 | cefazolin | RxNorm |
| 43009083 | cefroxadine | RxNorm Extension |
| 43008994 | ceftezole sodium | RxNorm Extension |
| 1786621 | cephalexin | RxNorm |
| 19052683 | cephaloridine | RxNorm |
| 19086759 | cephalothin | RxNorm |
| 19086790 | cephapirin | RxNorm |
| 1786842 | cephradine | RxNorm |
| 990069 | chloramphenicol | RxNorm |
| 997881 | clindamycin | RxNorm |
| 36863501 | clometocillin | RxNorm Extension |
| 1800835 | cloxacillin | RxNorm |
| 1724666 | dicloxacillin | RxNorm |
| 1738521 | doxycycline | RxNorm |
| 36851057 | epicillin | RxNorm Extension |
| 19054936 | floxacillin | RxNorm |
| 45892419 | gentamicin | RxNorm |
| 19069006 | hetacillin | RxNorm |
| 19072054 | methampicillin | RxNorm |
| 1361385 | methicillin | RxNorm |
| 1707164 | metronidazole | RxNorm |
| 1713930 | nafcillin | RxNorm |
| 40798981 | nifurtoinol | RxNorm Extension |
| 920293 | nitrofurantoin | RxNorm |
| 19024197 | ornidazole | RxNorm |
| 1724703 | oxacillin | RxNorm |
| 36857825 | penamecillin | RxNorm Extension |
| 1728416 | penicillin G | RxNorm |
| 1729720 | penicillin V | RxNorm |
| 19047071 | pivampicillin | RxNorm |
| 19096054 | propicillin | RxNorm |
| 19037983 | secnidazole | RxNorm |
| 1701651 | spectinomycin | RxNorm |
| 1836241 | sulbactam | RxNorm |
| 1836391 | sulfadiazine | RxNorm |
| 19000818 | sulfadimethoxine | RxNorm |
| 19136423 | sulfalene | RxNorm |
| 36856579 | sulfamazone | RxNorm Extension |
| 19136426 | sulfamerazine | RxNorm |
| 19136429 | sulfamethazine | RxNorm |
| 1036425 | sulfamethizole | RxNorm |
| 1836430 | sulfamethoxazole | RxNorm |
| 19000820 | sulfamethoxypyridazine | RxNorm |
| 36854873 | sulfametomidine | RxNorm Extension |
| 40799118 | sulfametoxydiazine | RxNorm Extension |
| 19040624 | sulfametrole | RxNorm |
| 19136481 | sulfamoxole | RxNorm |
| 1036475 | sulfanilamide | RxNorm |
| 40799120 | sulfaperin | RxNorm Extension |
| 40799121 | sulfaphenazole | RxNorm Extension |
| 19136493 | sulfapyridine | RxNorm |
| 1036487 | sulfathiazole | RxNorm |
| 19000821 | sulfisomidine | RxNorm |
| 1836503 | sulfisoxazole | RxNorm |
| 43009009 | sultamicillin | RxNorm Extension |
| 19002077 | talampicillin | RxNorm |
| 1836948 | tetracycline | RxNorm |
| 19137362 | thiamphenicol | RxNorm |
| 1702559 | tinidazole | RxNorm |
| 1705674 | trimethoprim | RxNorm |

### Table S3: Conditions of interest and their ancestor concepts.

| **Condition of Interest** | **Ancestor Concept IDs** | **Ancestor Concept Names** | **Notes** |
| --- | --- | --- | --- |
| Cardiac infection | 314383 | Myocarditis |  |
|  | 4138837 | Pericarditis |  |
|  | 441589 | Endocarditis |  |
| Care-related infection | 4193161 | Disorder following clinical procedure |  |
|  | 318445 | Post cardiac operation functional disturbance |  |
|  | 4123283 | Disorder of stoma |  |
|  | 43021974 | Complication associated with device |  |
|  | 4201387 | Tracheostomy present |  |
|  | 442019 | Complication of procedure |  |
| Ear infection | 4044878 | Infection of ear |  |
|  | 380731 | Otitis externa |  |
| Exacerbation of COPD | 255573 | Chronic obstructive pulmonary disease |  |
|  | 257004 | Acute exacerbation of chronic obstructive pulmonary disease |  |
| Exacerbation of cystic fibrosis | 441267 | Cystic fibrosis |  |
|  | 44808532 | Exacerbation of cystic fibrosis |  |
| Eye infection | 4134613 | Eye infection |  |
|  | 37160823 | Infection of eyelid caused by Mycobacterium leprae |  |
| GI infection | 37396146 | Gastrointestinal infection |  |
|  | 44783254 | Infection of masticator space |  |
|  | 198678 | Intestinal infectious disease |  |
|  | 198337 | Infectious diarrheal disease |  |
| Infection caused by antimicrobial resistant bacteria | 4249827 | Infection caused by antimicrobial resistant bacteria |  |
|  | 37017452 | Drug resistance to antibacterial agent |  |
|  | 44806682 | Infection resistant to multiple antibiotics |  |
| Lower respiratory tract infection | 4175297 | Lower respiratory tract infection |  |
|  | 4133224 | Lobar pneumonia |  |
|  | 618954 | Exacerbation of bronchiectasis caused by infection |  |
| Neutropenia and agranulocytosis | 320073 | Neutropenia |  |
|  | 440689 | Agranulocytosis |  |
| Other infection | 432545 | Bacterial infectious disease | Excluding concepts found in other conditions of interest |
| Sepsis and septic shock | 132797 | Sepsis |  |
|  | 196236 | Septic shock |  |
| Signs and symptoms | 201965 | Shock |  |
|  | 437663 | Fever |  |
|  | 254761 | Cough |  |
|  | 4305080 | Abnormal breathing |  |
|  | 31967 | Nausea |  |
|  | 441408 | Vomiting |  |
|  | 196523 | Diarrhoea |  |
|  | 254061 | Pleural effusion |  |
|  | 197672 | Urinary incontinence |  |
|  | 4103189 | Finding of heart rate | Excluding normal heart rate findings |
|  | 373995 | Delirium |  |
|  | 433595 | Oedema |  |
|  | 321689 | Apnoea |  |
|  | 253321 | Stridor |  |
|  | 200528 | Ascites |  |
|  | 435517 | Acidosis |  |
|  | 4128820 | Feeding finding |  |
|  | 435515 | Hypo-osmolality and or hyponatremia |  |
|  | 4214962 | Blood pressure finding | Excluding normal blood pressure findings |
|  | 44784217 | Cardiac arrhythmia |  |
| Skin infection | 201093 | Infection of skin and/or subcutaneous tissue |  |
|  | 141095 | Acne |  |
| Upper respiratory tract infection | 4181583 | Upper respiratory infection |  |
|  | 4234533 | Tonsillitis |  |
| Urinary tract infection | 81902 | Urinary tract infectious disease |  |

### Table S4: Patient characteristics for the top five antibiotics in Barts

| **Variable name** | **Variable level** | **Estimate name** | **Piperacillin / Tazobactam** | **Clarithromycin** | **Ciprofloxacin** | **Ceftriaxone** | **Meropenem** |
| --- | --- | --- | --- | --- | --- | --- | --- |
| Number  records |  | N | 19,976 | 11,794 | 11,359 | 10,841 | 7,417 |
| Number  subjects |  | N | 16,392 | 10,562 | 9,761 | 10,250 | 6,276 |
| Cohort start  date |  | Range | 2022-01-01  to  2025-05-04 | 2022-01-01  to  2025-05-03 | 2022-01-01  to  2025-05-04 | 2022-01-01  to  2025-05-04 | 2022-01-01  to  2025-05-03 |
| Age |  | Median  [Q25 –  Q75] | 65 [50 - 77] | 65 [44 - 79] | 62 [43 - 76] | 38 [19 - 60] | 61 [44 - 74] |
|  |  | Range | 0 to 113 | 0 to 114 | 0 to 113 | 0 to 114 | 0 to 102 |
| Sex | Female | N (%) | 8,554  (42.82%) | 5,981  (50.71%) | 5,644  (49.69%) | 4,993  (46.06%) | 3,212  (43.31%) |
|  | Male | N (%) | 11,422  (57.18%) | 5,813  (49.29%) | 5,715  (50.31%) | 5,848  (53.94%) | 4,205  (56.69%) |

### Table S5: Patient characteristics for the top five antibiotics in GOSH

| **Variable name** | **Variable level** | **Estimate name** | **Piperacillin / Tazobactam** | **Teicoplanin** | **Vancomycin** | **Ciprofloxacin** | **Meropenem** |
| --- | --- | --- | --- | --- | --- | --- | --- |
| Number  records |  | N | 4,227 | 2,025 | 1,757 | 1,571 | 1,527 |
| Number  subjects |  | N | 3,143 | 1,635 | 1,367 | 1,204 | 1,173 |
| Cohort  start date |  | Range | 2022-01-01  to  2025-02-17 | 2022-01-04  to  2025-02-17 | 2022-01-02  to  2025-02-16 | 2022-01-02  to  2025-02-17 | 2022-01-03  to  2025-02-17 |
| Age |  | Median  [Q25 –  Q75] | 2 [0 - 8] | 4 [0 - 11] | 1 [0 - 7] | 4 [1 - 10] | 2 [0 - 7] |
|  |  | Range | 0 to 17 | 0 to 17 | 0 to 17 | 0 to 17 | 0 to 17 |
| Sex | Female | N (%) | 1,772  (41.92%) | 879  (43.41%) | 757  (43.08%) | 692  (44.05%) | 644  (42.17%) |
|  | Male | N (%) | 2,455  (58.08%) | 1,146  (56.59%) | 1,000  (56.92%) | 879  (55.95%) | 883  (57.83%) |

### Table S6: Patient characteristics for the top five antibiotics in Lancashire

| **Variable name** | **Variable level** | **Estimate name** | **Cefuroxime** | **Clarithromycin** | **Piperacillin / Tazobactam** | **Ciprofloxacin** | **Levofloxacin** |
| --- | --- | --- | --- | --- | --- | --- | --- |
| Number  records |  | N | 10,801 | 9,870 | 6,642 | 2,400 | 2,314 |
| Number  subjects |  | N | 9,634 | 8,281 | 5,857 | 2,177 | 1,984 |
| Cohort  start date |  | Range | 2022-01-01  to  2024-12-30 | 2022-01-01  to  2025-01-01 | 2022-01-01  to  2024-12-31 | 2022-01-01  to  2024-12-29 | 2022-01-02  to  2024-12-27 |
| Age |  | Median  [Q25 –  Q75] | 69 [50 - 80] | 75 [62 - 84] | 72 [60 - 81] | 68 [55 - 78] | 76 [62 - 84] |
|  |  | Range | 0 to 104 | 0 to 104 | 0 to 104 | 1 to 102 | 7 to 101 |
| Sex | Female | N (%) | 5,475  (50.69%) | 4,655  (47.16%) | 2,843  (42.80%) | 1,184  (49.33%) | 1,222  (52.81%) |
|  | Male | N (%) | 5,326  (49.31%) | 5,215  (52.84%) | 3,799  (57.20%) | 1,216  (50.67%) | 1,092  (47.19%) |

### Table S7: Patient characteristics for the top five antibiotics in Leeds

| **Variable name** | **Variable level** | **Estimate name** | **Piperacillin / Tazobactam** | **Clarithromycin** | **Cefuroxime** | **Ciprofloxacin** | **Teicoplanin** |
| --- | --- | --- | --- | --- | --- | --- | --- |
| Number  records |  | N | 26,365 | 11,567 | 11,270 | 10,043 | 9,608 |
| Number  subjects |  | N | 20,460 | 10,318 | 10,412 | 8,661 | 8,401 |
| Cohort  start date |  | Range | 2022-01-01  to  2025-03-18 | 2022-01-01  to  2025-03-19 | 2022-01-01  to  2025-03-19 | 2022-01-01  to  2025-03-19 | 2022-01-01  to  2025-03-18 |
| Age |  | Median  [Q25 –  Q75] | 72 [57 - 81] | 72 [54 - 82] | 40 [28 - 60] | 64 [44 - 77] | 61 [42 - 75] |
|  |  | Range | 0 to 108 | 0 to 108 | 0 to 100 | 0 to 102 | 0 to 105 |
| Sex | Female | N (%) | 11,562  (43.85%) | 5,522  (47.74%) | 7,178  (63.69%) | 4,845  (48.24%) | 4,569  (47.55%) |
|  | Male | N (%) | 14,803  (56.15%) | 6,045  (52.26%) | 4,092  (36.31%) | 5,198  (51.76%) | 5,039  (52.45%) |

### Table S8: Patient characteristics for the top five antibiotics in UCLH

| **Variable name** | **Variable level** | **Estimate name** | **Cefuroxime** | **Ciprofloxacin** | **Piperacillin / Tazobactam** | **Clarithromycin** | **Teicoplanin** |
| --- | --- | --- | --- | --- | --- | --- | --- |
| Number  records |  | N | 44,228 | 19,251 | 10,450 | 9,842 | 5,547 |
| Number  subjects |  | N | 39,445 | 16,506 | 7,995 | 8,759 | 4,887 |
| Cohort  start date |  | Range | 2022-01-01  to  2025-05-12 | 2022-01-01  to  2025-05-12 | 2022-01-01  to  2025-05-09 | 2022-01-01  to  2025-05-12 | 2022-01-01  to  2025-05-09 |
| Age |  | Median  [Q25 –  Q75] | 50 [35 - 67] | 52 [32 - 67] | 59 [41 - 71] | 57 [36 - 71] | 57 [37 - 70] |
|  |  | Range | 0 to 107 | 0 to 101 | 0 to 104 | 0 to 105 | 0 to 104 |
| Sex | Female | N (%) | 27,009  (61.07%) | 9,688  (50.32%) | 4,541  (43.45%) | 5,175  (52.58%) | 2,995  (53.99%) |
|  | Male | N (%) | 17,219  (38.93%) | 9,563  (49.68%) | 5,909  (56.55%) | 4,667  (47.42%) | 2,552  (46.01%) |

| **Variable name** | **Variable level** | **Estimate name** | **Neomycin** | **Fusidate** | **Ciprofloxacin** | **Erythromycin** | **Clarithromycin** |
| --- | --- | --- | --- | --- | --- | --- | --- |
| Number  records |  | N | 4,477,598 | 2,029,304 | 1,561,067 | 1,295,225 | 769,070 |
| Number  subjects |  | N | 2,627,866 | 1,446,531 | 1,019,792 | 722,514 | 537,083 |
| Cohort  start date |  | Range | 2012-01-01  to  2024-12-18 | 2012-01-01  to  2024-12-17 | 2012-01-01   to  2024-12-18 | 2012-01-01   to  2024-12-18 | 2012-01-01   to  2024-12-17 |
| Age |  | Median  [Q25 –  Q75] | 49 [28 - 66] | 44 [11 - 68] | 59 [41 - 74] | 16 [6 - 31] | 6 [3 - 13] |
|  |  | Range | 0 to 111 | 0 to 111 | 0 to 109 | 0 to 111 | 0 to 112 |
| Sex | Female | N (%) | 2,535,489 (56.63%) | 1,114,817  (54.94%) | 769,680  (49.30%) | 805,653  (62.20%) | 399,320  (51.92%) |
|  | Male | N (%) | 1,942,109 (43.37%) | 914,487 (45.06%) | 791,387  (50.70%) | 489,572   (37.80%) | 369,750  (48.08%) |

### Table S9: Patient characteristics for the top five antibiotics in CPRD Aurum

### Table S10: Patient characteristics for the top five antibiotics in DataLoch

| **Variable name** | **Variable level** | **Estimate name** | **Clarithromycin** | **Fusidate** | **Neomycin** | **Oxytetracycline** | **Erythromycin** |
| --- | --- | --- | --- | --- | --- | --- | --- |
| Number  records |  | N | 394,394 | 390,456 | 353,001 | 178,584 | 136,037 |
| Number  subjects |  | N | 209,695 | 237,399 | 173,806 | 65,386 | 60,640 |
| Cohort  start date |  | Range | 2012-01-31   to  2025-03-31 | 2012-01-31  to  2025-03-31 | 2012-01-31 to  2025-03-31 | 2012-01-31   to  2025-03-31 | 2012-01-31  to  2025-03-31 |
| Age |  | Median  [Q25 –  Q75] | 50 [31 - 66] | 38 [14 - 62] | 51 [31 - 66] | 50 [31 - 65] | 30 [18 - 48] |
|  |  | Range | 0 to 108 | 0 to 107 | 0 to 105 | 0 to 105 | 0 to 107 |
| Sex | Female | N (%) | 244,700  (62.04%) | 215,991   (55.32%) | 206,292  (58.44%) | 92,335  (51.70%) | 87,630  (64.42%) |
|  | Male | N (%) | 149,694  (37.96%) | 174,465  (44.68%) | 146,709  (41.56%) | 86,249  (48.30%) | 48,407  (35.58%) |

### Figure S1: Quarterly crude incidence rates of the top five Watch antibiotic used in Barts, stratified by sex.

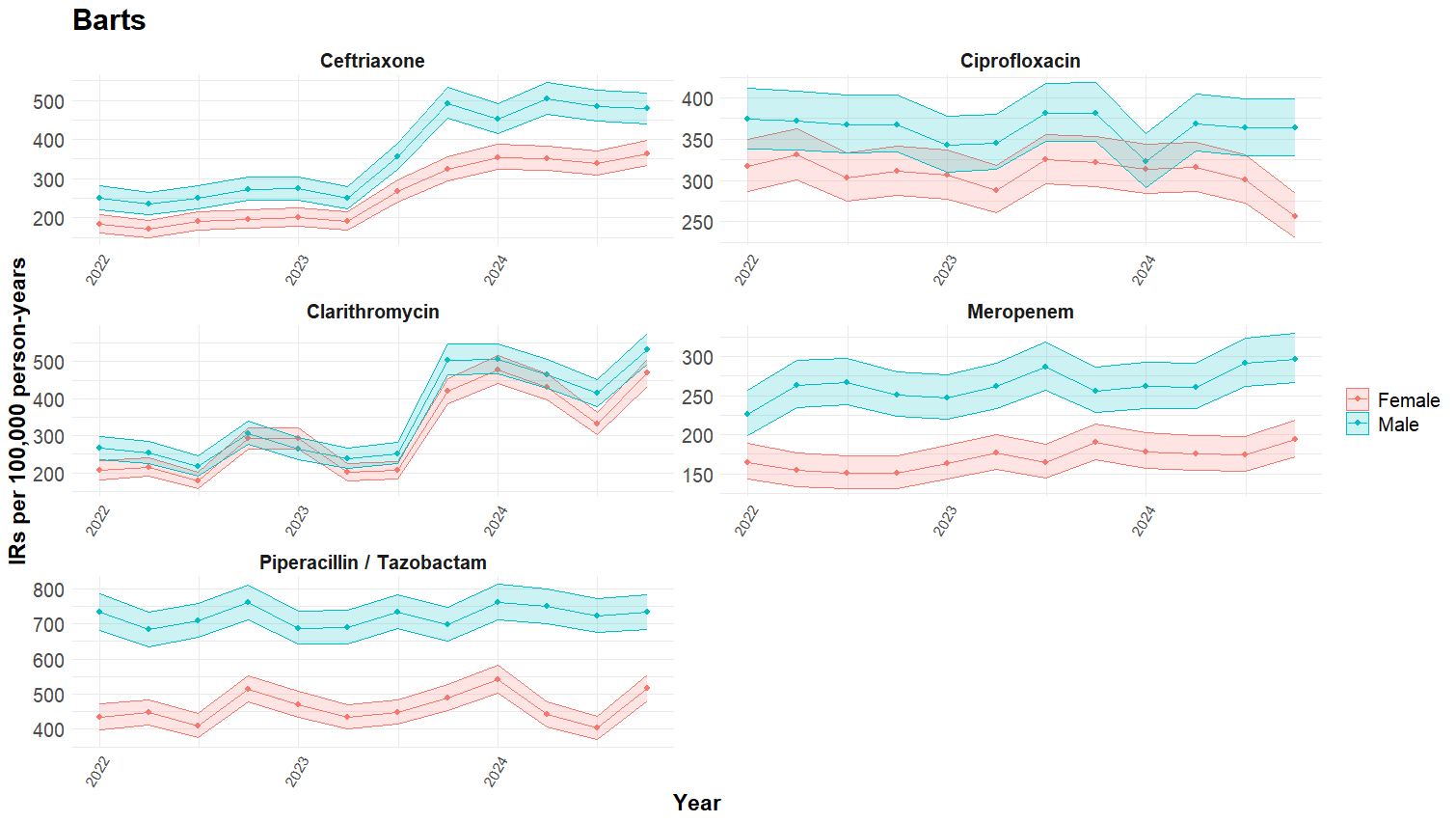

### Figure S2: Quarterly crude incidence rates of the top five Watch antibiotic used in GOSH, stratified by sex.

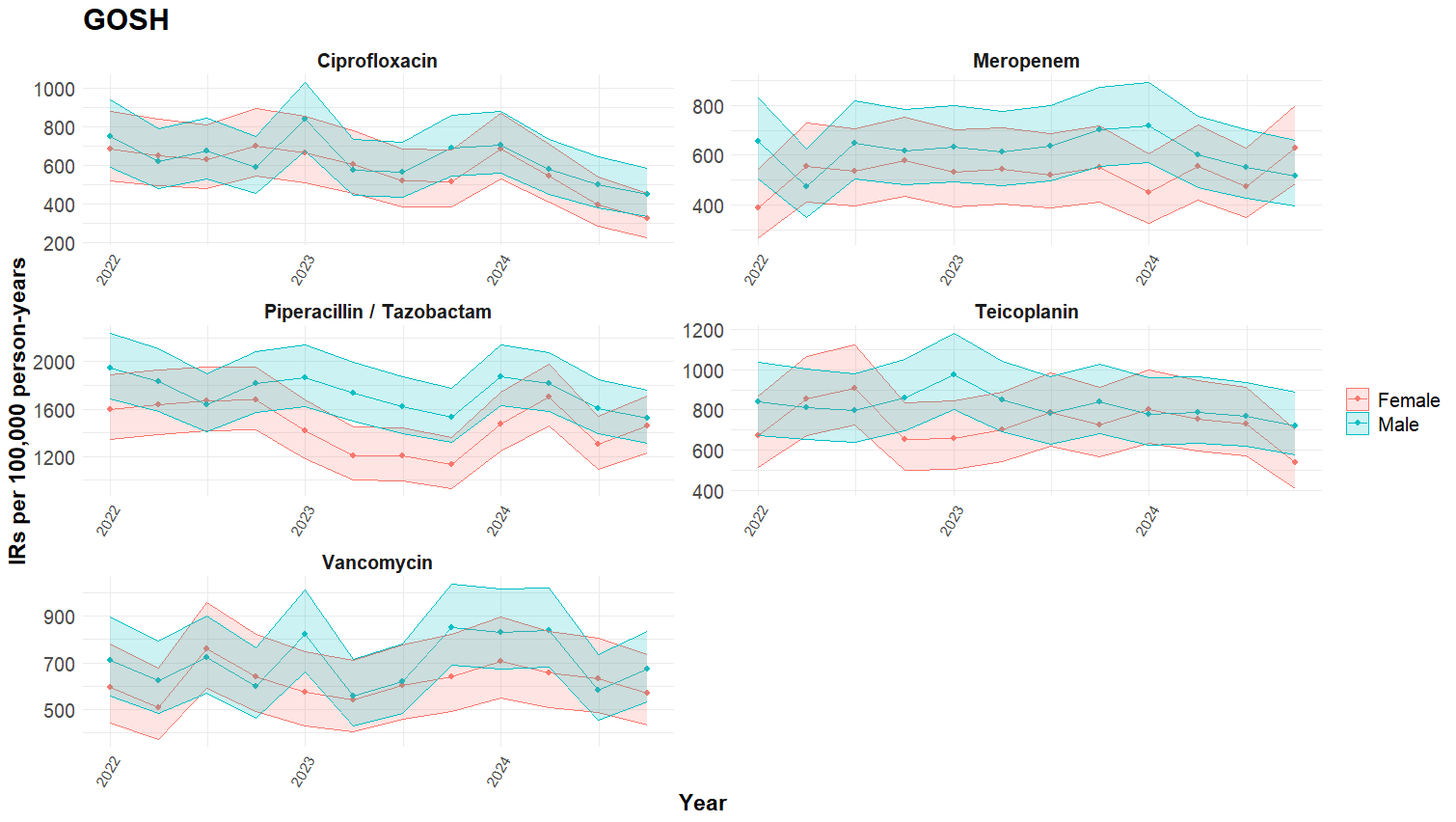

### Figure S3: Quarterly crude incidence rates of the top five Watch antibiotic used in Lancashire, stratified by sex.

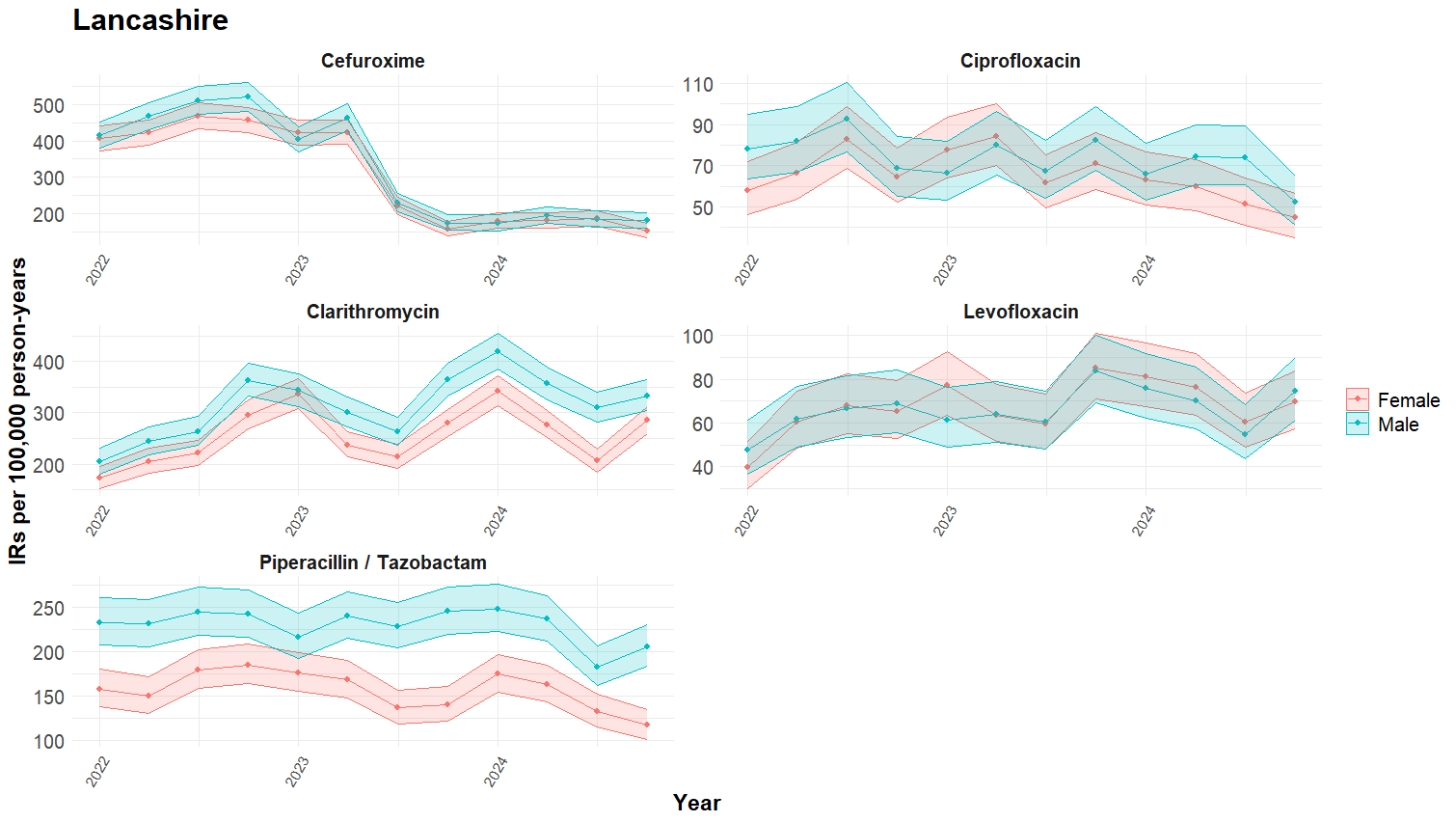

### Figure S4: Quarterly crude incidence rates of the top five Watch antibiotic used in Leeds, stratified by sex.

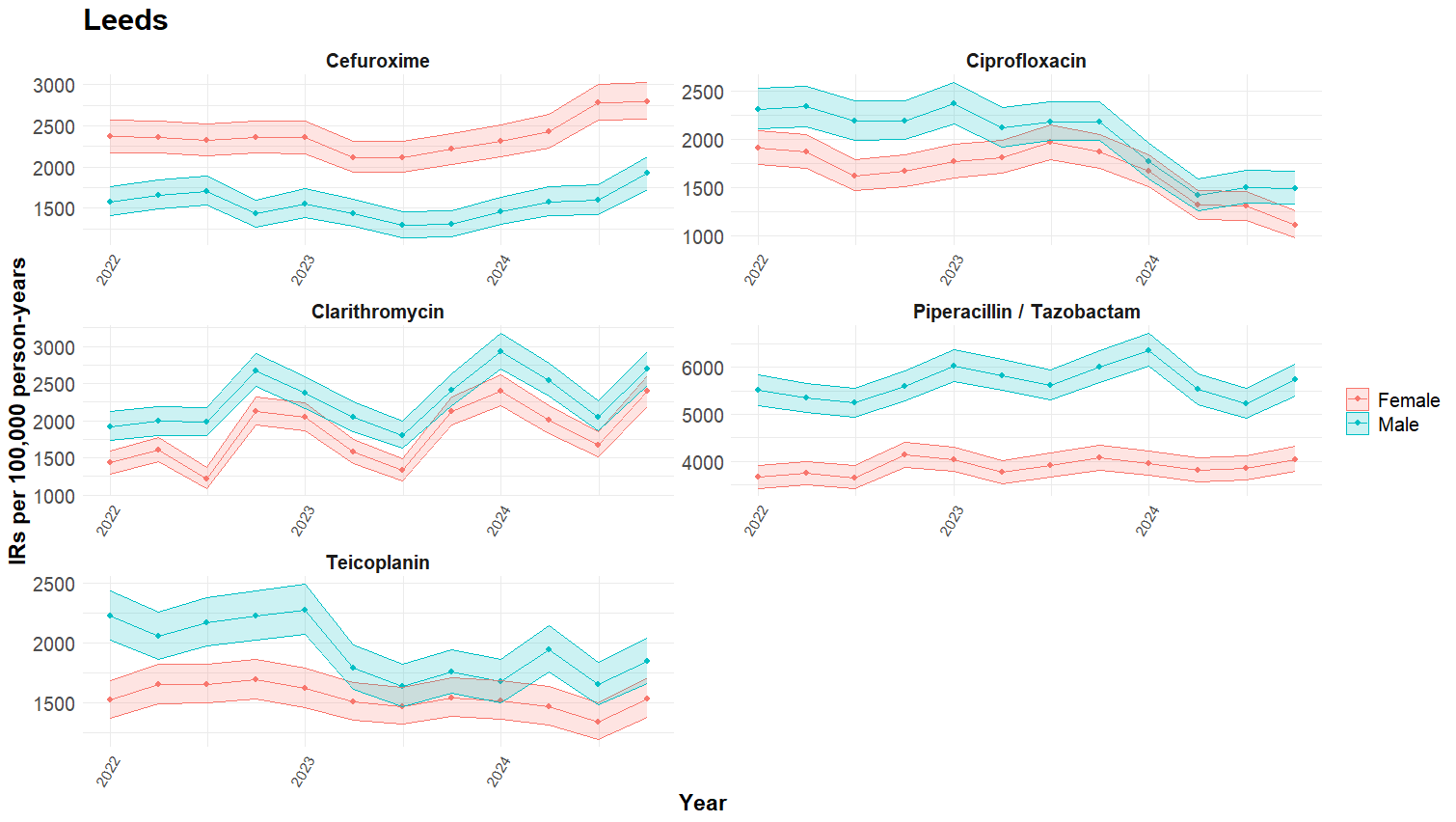

### Figure S5: Quarterly crude incidence rates of the top five Watch antibiotic used in UCLH, stratified by sex.

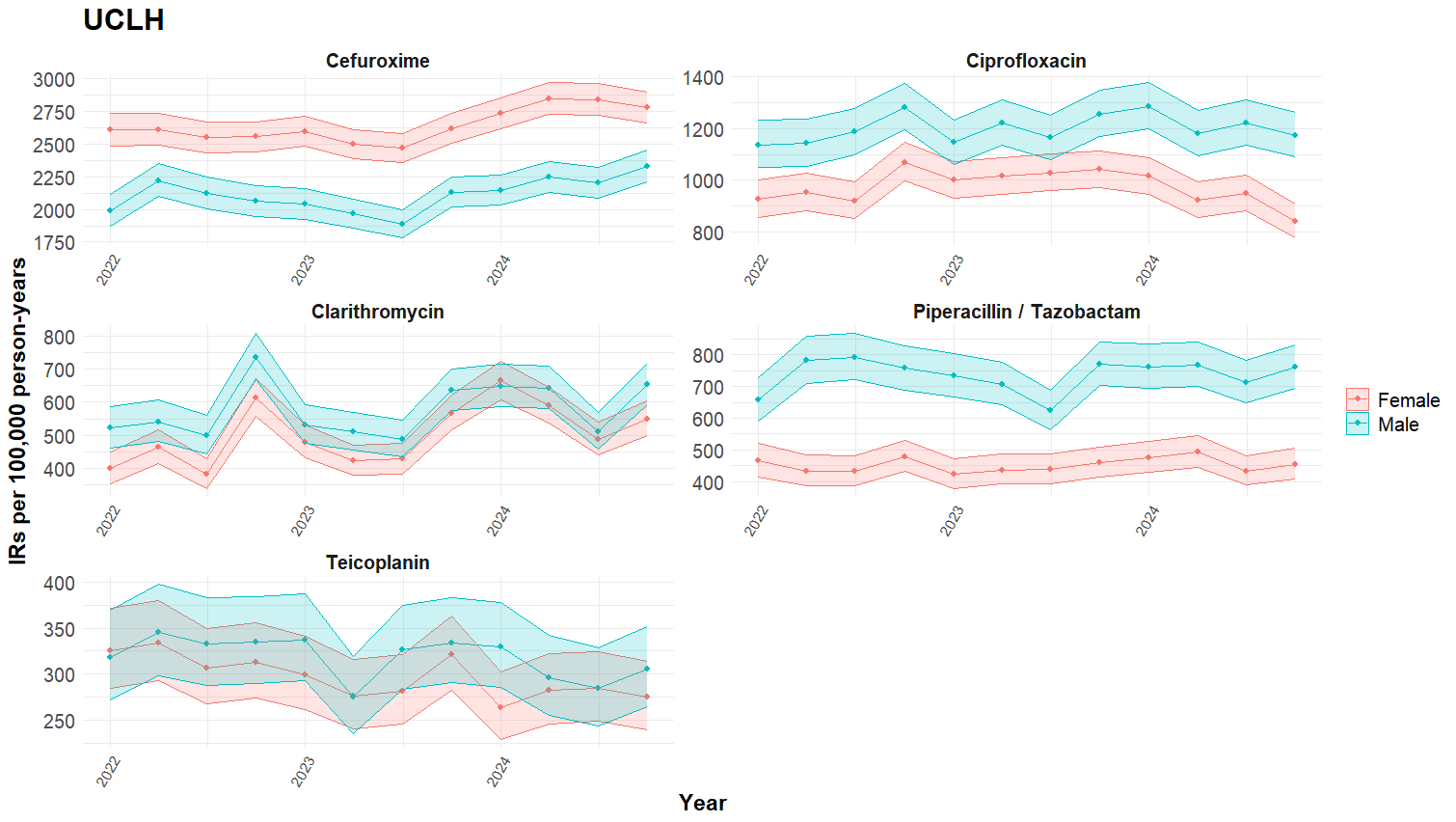

### Figure S6: Quarterly crude incidence rates of the top five Watch antibiotic used in CPRD Aurum, stratified by sex.

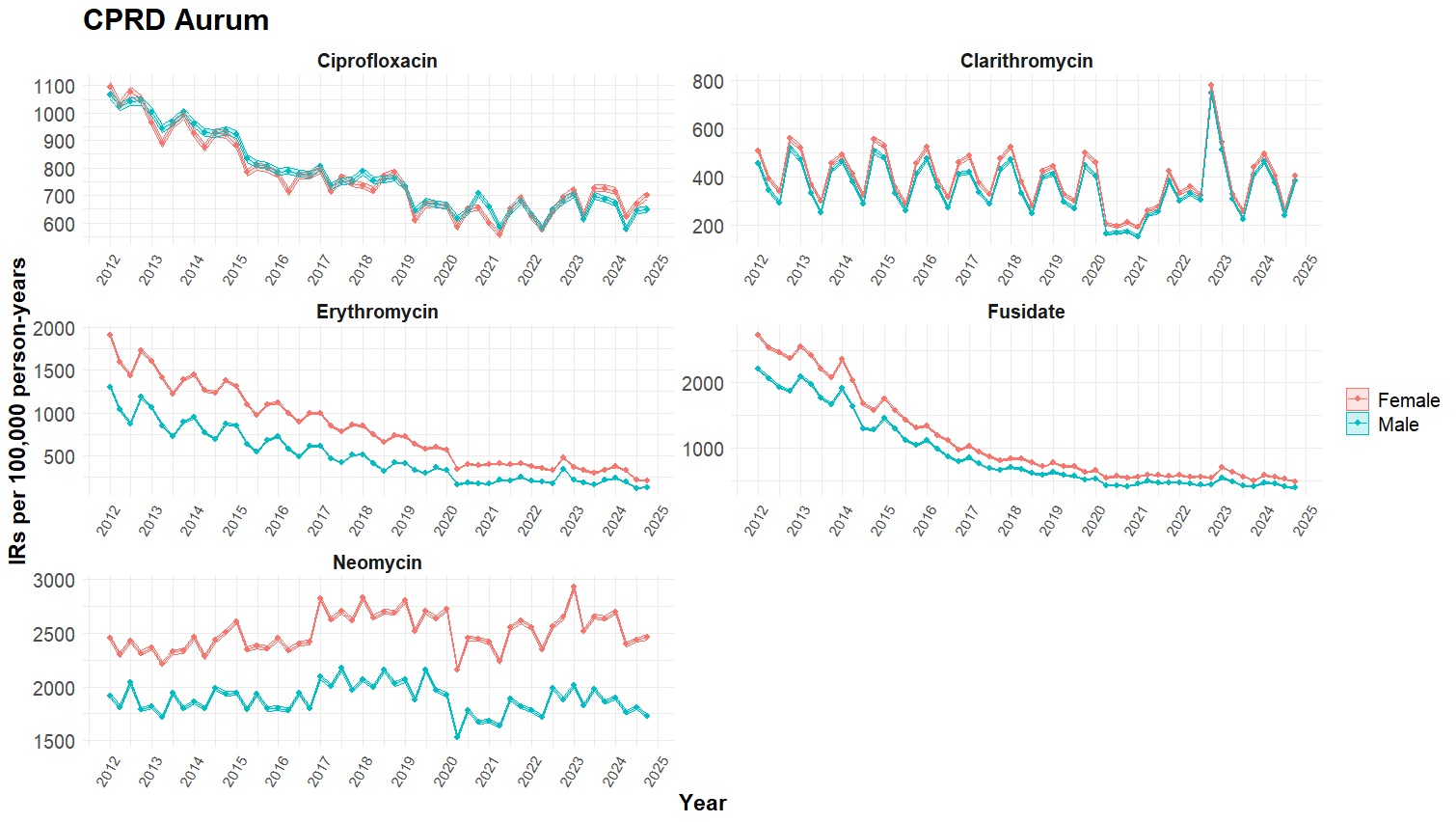

### Figure S7: Quarterly crude incidence rates of the top five Watch antibiotic used in DataLoch, stratified by sex.

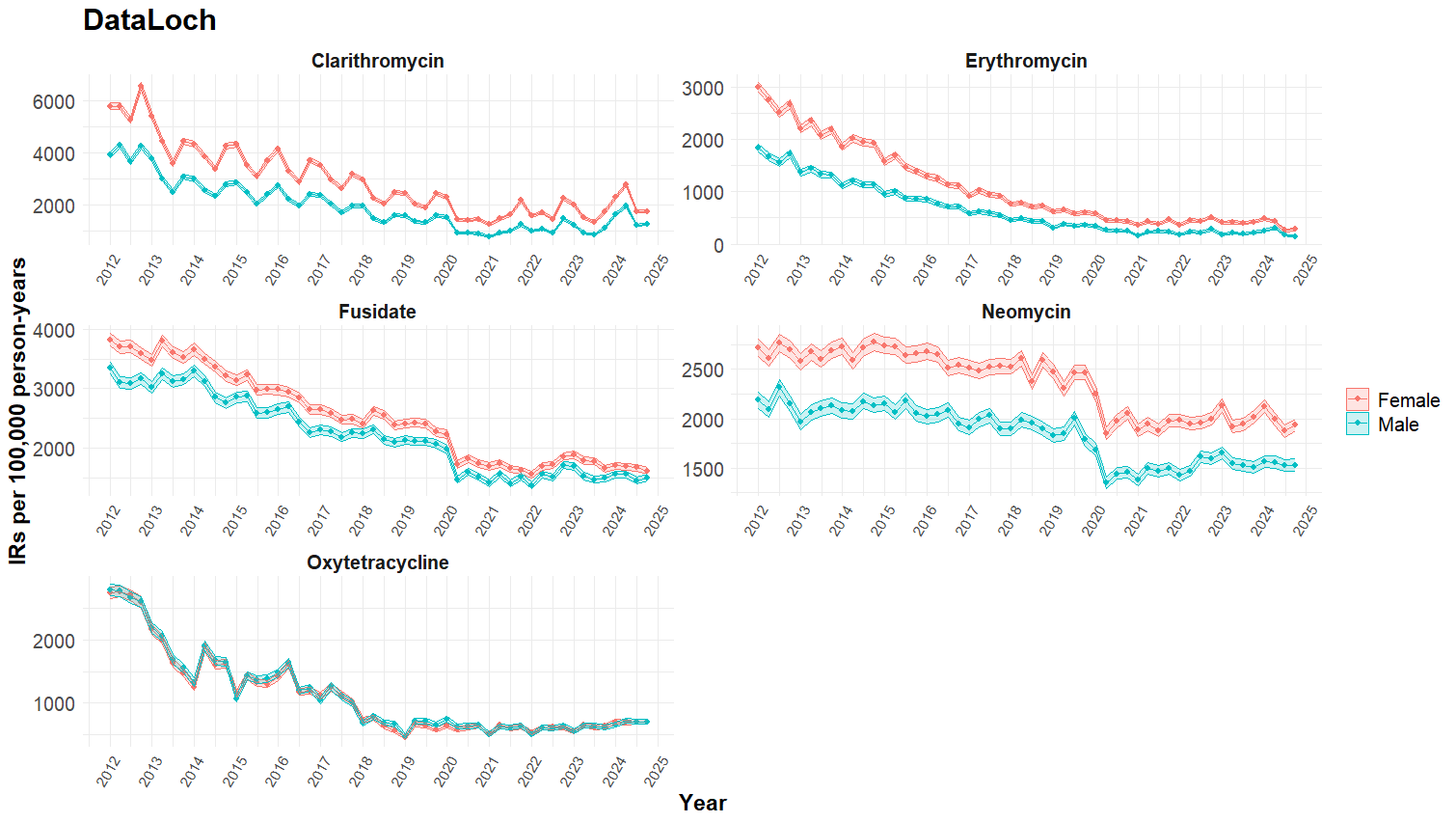

### Figure S8: Quarterly crude incidence rates of the top five Watch antibiotic used in Barts, stratified by broad age group.

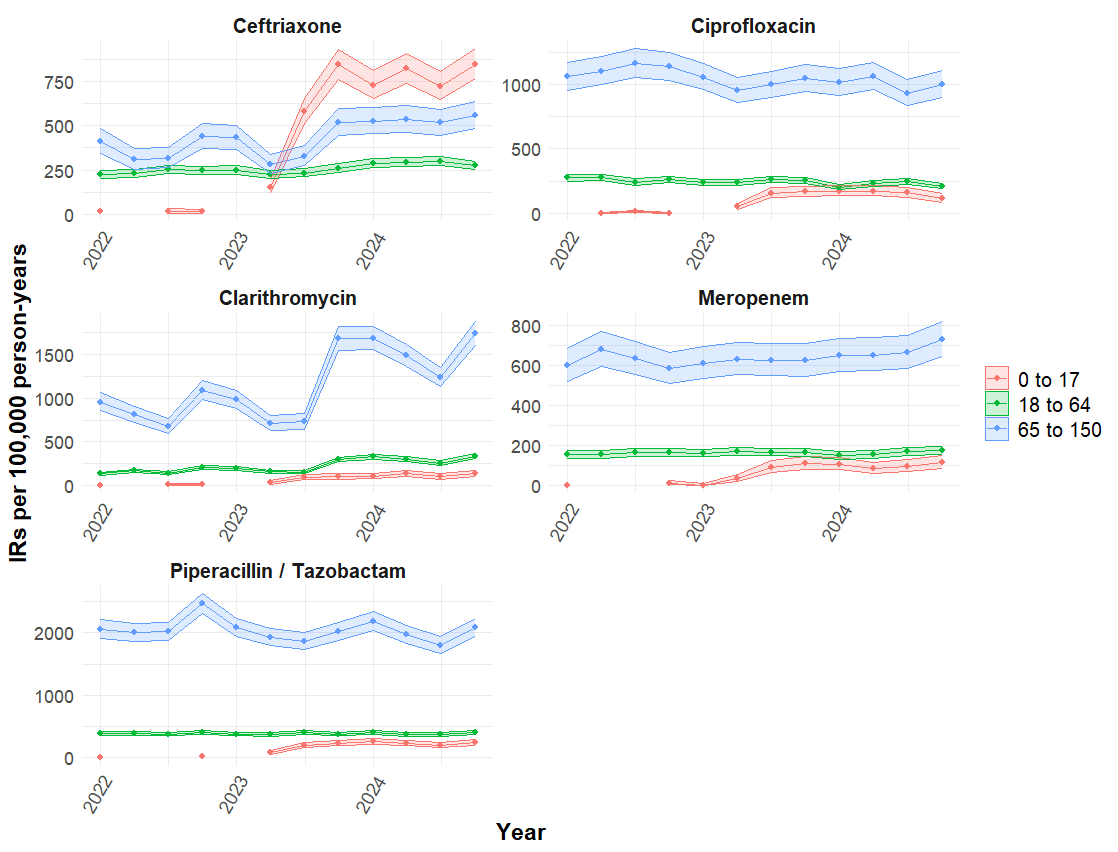

### Figure S9: Quarterly crude incidence rates of the top five Watch antibiotic used in Lancashire, stratified by broad age group.

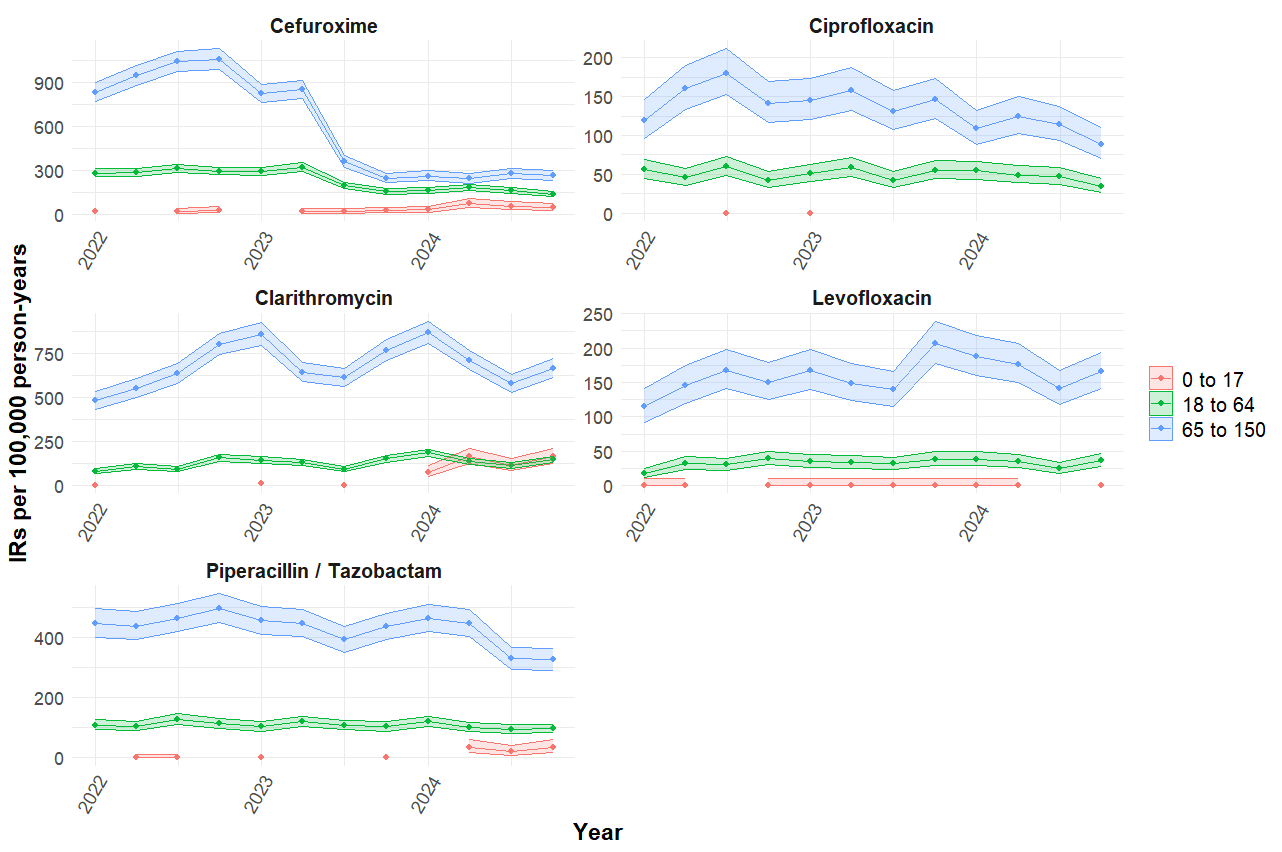

### Figure S10: Quarterly crude incidence rates of the top five Watch antibiotic used in Leeds, stratified by broad age group.

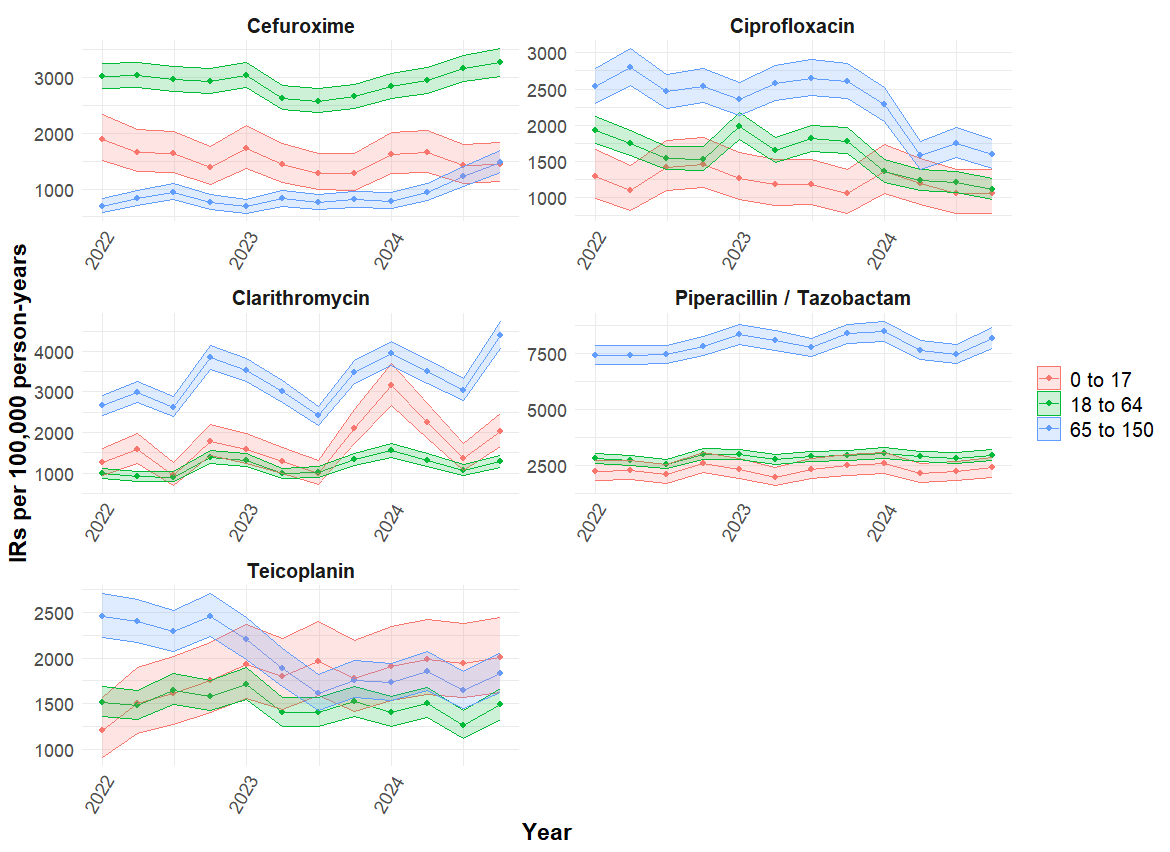

### Figure S11: Quarterly crude incidence rates of the top five Watch antibiotic used in UCLH, stratified by broad age group.

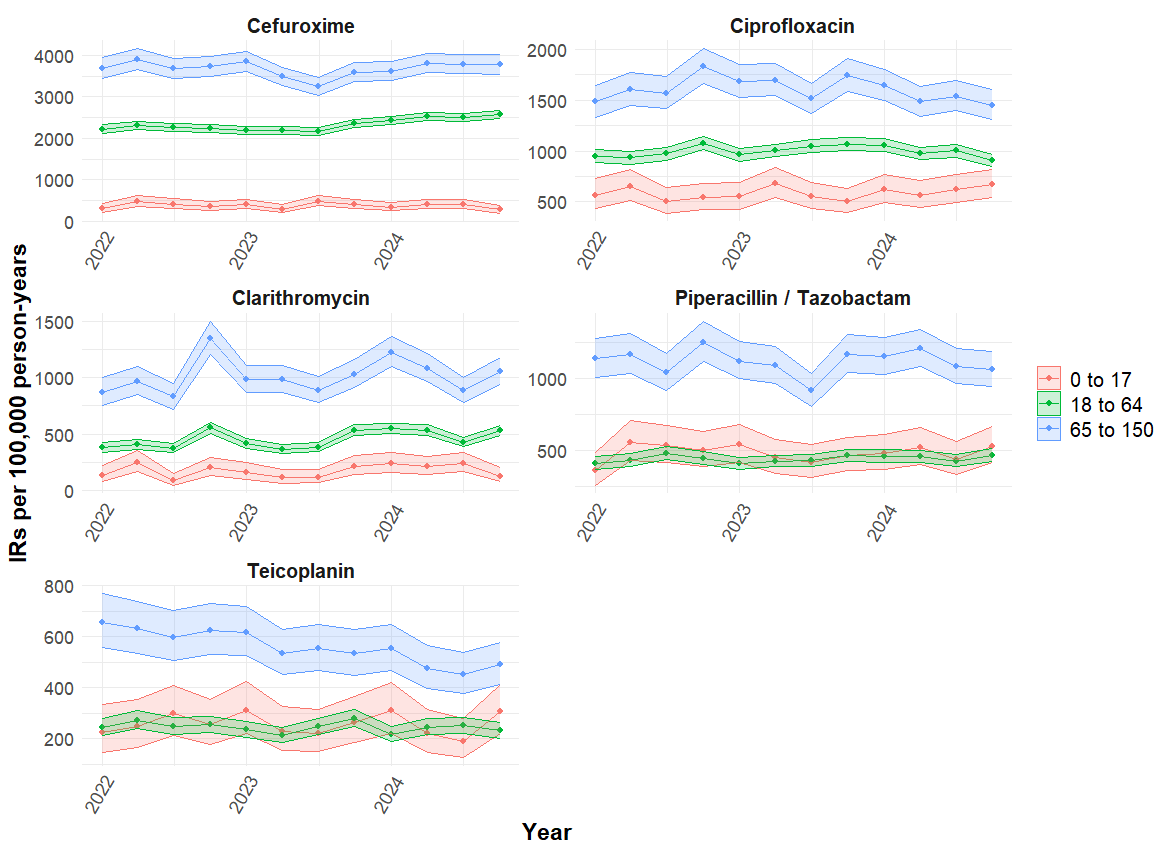

### Figure S12: Quarterly crude incidence rates of the top five Watch antibiotic used in CPRD Aurum, stratified by broad age group.

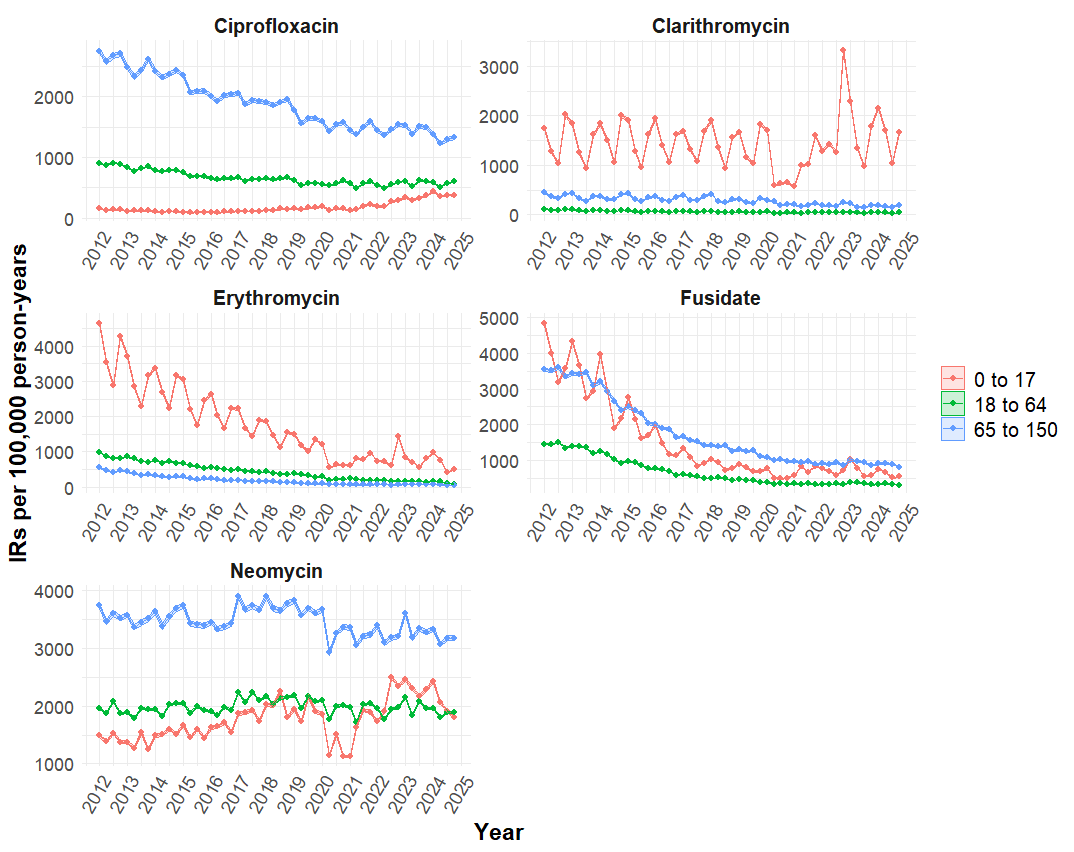

### Figure S13: Quarterly crude incidence rates of the top five Watch antibiotic used in DataLoch, stratified by broad age group.

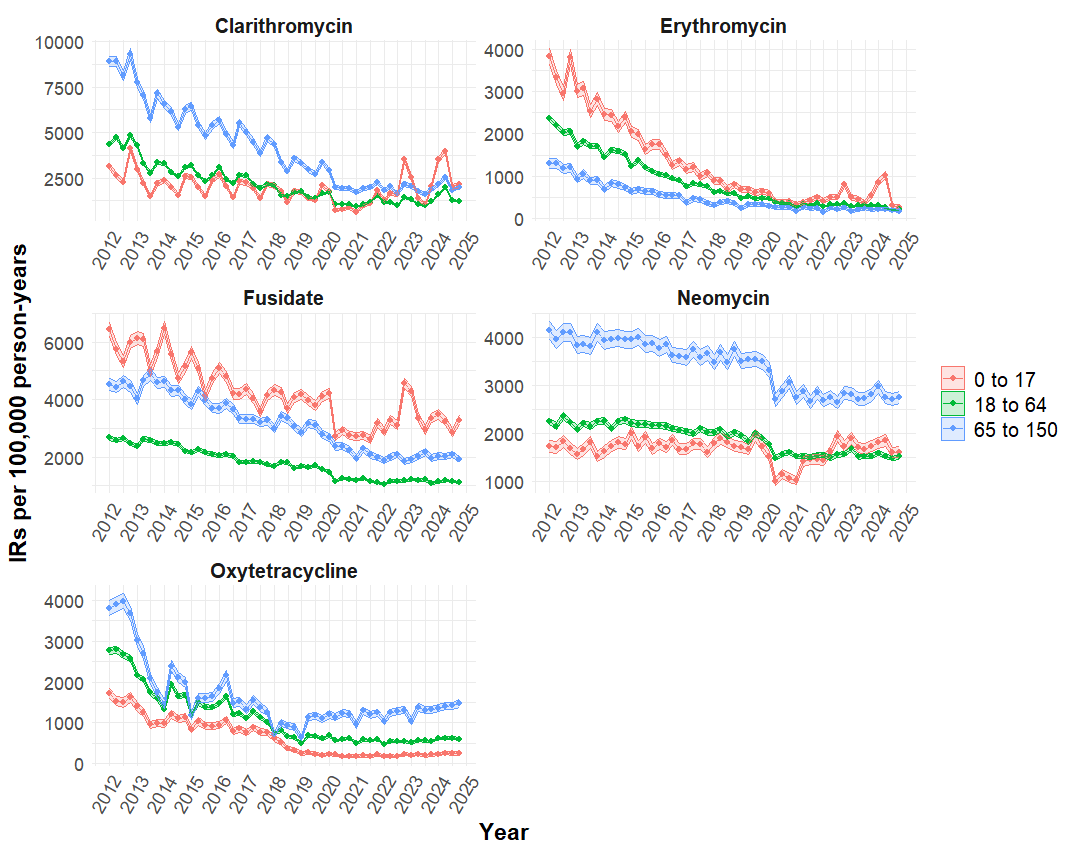
